# Electrophysiologically-defined excitation-inhibition autism neurosubtypes

**DOI:** 10.1101/2023.11.22.23298729

**Authors:** Natasha Bertelsen, Gabriele Mancini, David Sastre-Yagüe, Andrea Vitale, Gabriel Matías Lorenz, Simone Blanco Malerba, Dimitris Bolis, Veronica Mandelli, Pablo Martínez-Cañada, Elena Maria Busuoli, Sara Danieli, Maria Facchini, Oscar Joseph Lega, Ines Severino, Margherita Raffaelli, Arianna Bentenuto, Paola Venuti, Nicola Palumbo, Katia Pedrolli, Corrado Barone, Stefano Calzolari, Emanuela Contardo, Alessandro Gozzi, Stefano Panzeri, Michael V. Lombardo

## Abstract

Neurophysiological excitation versus inhibition (E:I) imbalance has long been theorized as one of the primary neurobiological explanations behind autism. However, progress applying this theory to most autistic individuals has been impeded by limitations in understanding of how non-invasive electrophysiological data (e.g., EEG) can be translationally used to pinpoint underlying E:I mechanisms. Using *in-silico* modeling alongside *in-vivo* animal validations, we show that a fractal component measured by the Hurst exponent (H) tracks with single neuron excitability, while γ oscillations tracks with the ratio of E versus I synaptic conductances. Both H and γ are complementary for predicting overall network excitability. In human EEG data we find two autism neurosubtypes characterized by opposing profiles of H and γ and different large-scale brain-behavioral relationships affecting language, cognition, and other co-occurring neuropsychiatric issues. This work establishes that non-invasively measured electrophysiological signals can track with different aspects of E:I balance and that autism is composed of opposing E:I neurosubtypes.

## Background

Imbalance between neurophysiological excitation (E) and inhibition (I) (henceforth referred to as ‘E:I’ imbalance) has long been theorized to be one of the primary neurobiological explanations behind autism^1^. While the theory has been highly influential, it rests on evidence primarily gleaned from animal model research on known high-impact genomic mechanisms linked to autism^2,3^. Nearly all of these genomic causes are rare in frequency both within the general population, but also within autism^4^. Furthermore, the directionality (i.e. increased versus decreased E:I balance) and interpretation (e.g., initial cause, homeostatic compensation)^2,3,5^ of observed E:I imbalance in such rare genomic causes can markedly vary. Thus, a clear roadblock exists with regard to generalizing heterogeneous evidence of E:I imbalance from such rare genomic causes to the vast majority of autistic individuals whose genetic etiologies are otherwise unknown (i.e. idiopathic autism). Sohal and Rubenstein^3^ proposed that the next steps in moving forward are towards identifying more individualized biomarkers of different types of E:I imbalance which might lead towards differential explanations behind heterogeneity in behavioral phenotypes and/or differential responses to treatment. In this work, we take these next steps by first enhancing our understanding of how we can decode underlying E:I mechanisms via *in-vivo* measurement of electrophysiological time-series features from electroencephalography (EEG) data. With a more precise mechanistic understanding of how EEG features may relate to different aspects of E:I balance, we then utilize such EEG E:I biomarkers to stratify autism into ‘neurosubtypes’ and test for different brain-behavioral relationships.

A prominent challenge in linking electrophysiological data (e.g., local field potentials, LFP, or EEG) to concepts like E:I balance lies in the fact that while such data can be decomposed into a plethora of different time-series features (e.g., ^6–9)^, it remains unclear how such features may capture disparate underlying E:I-relevant mechanisms. Thus, a key translational goal is to better connect how *in-vivo* electrophysiological time-series features allow for decoding specific types of underlying E:I-relevant mechanisms. One strategy for making progress is to first generate predictions via *in-silico* simulations of recurrently connected excitatory (E) and inhibitory (I) neuronal populations while manipulating ground truth underlying E:I parameters, and then to follow such work with experiments where E or I can be manipulated *in-vivo* and empirically tested. We thus start with *in-silico* manipulations of key E:I mechanisms: single neuron excitability and ratio of synaptic E:I conductances. We then identify specific electrophysiological time-series features that track with these key E:I mechanisms and use them as biomarkers for *in-vivo* experiments where E or I are manipulated chemogenetically in mice.

Of the many possible candidate EEG time-series features, we focus on two prominent features with initial links to E:I mechanisms - that is, the aperiodic or fractal component captured by the Hurst exponent (H) and a periodic oscillatory component captured by γ-band oscillations in the range of 30-50 Hz. Prior simulations of both coupled and uncoupled excitatory (E) and inhibitory (I) neuronal populations have shown that under contexts of low single neuron excitability, aperiodic/fractal features measured by the 1/f slope or H may be sensitive to underlying changes in the ratio of excitatory (E) versus inhibitory (I) synaptic conductances^10,11^. Here, we expand on this prior work by assessing how these aperiodic/fractal features behave under varying levels of single neuron excitability and synaptic E:I ratio, as well as by comparing how measures like H, 1/f slope, and total broadband power^12^ track underlying changes in E:I parameters. In addition to these aperiodic/fractal features, we also focus on γ-band oscillations within the range of 30-50 Hz. γ oscillations are a prominent result of E:I interactions^8,13–15^ and are highly relevant for understanding many aspects of brain function and dysfunction^3,16–18^. A key question we examine is whether H and γ features are sensitive to different underlying aspects of E:I balance. We also consider whether these different electrophysiological time-series features could be utilized in a complementary fashion to help predict emergent phenomena at the heart of understanding E:I balance - that is, decode overall ‘network excitability’, operationalized as the firing rate of the local neuronal network.

## Results

### In-silico modeling showing that H and γ power track different aspects of E:I balance

One primary goal of this work is to better understand how *in-vivo* electrophysiological neural time-series data (e.g., LFP and EEG) can be used to decode underlying E:I mechanisms. Towards this goal, we start by using *in-silico* modeling to derive predictions about how electrophysiological time-series features from LFP or EEG may decode such E:I mechanisms. Prior foundational work on this topic by Gao and colleagues^10^ simulated uncoupled E and I neuronal populations and linked aperiodic features like the 1/f slope of LFP power spectral density to the ratio of E versus I synaptic conductances^10^. One drawback of this approach is that the model is uncoupled and omits the fact that local E and I neuronal populations are recurrently connected. In Extended Data Fig. 1 we show that an uncoupled modeling approach is not sufficient for teasing apart effects of true E:I interactions (i.e. synaptic conductance ratio *g*) from independent changes in firing rates of E vs I neurons. Therefore, to achieve our goals we developed a model that includes recurrent interactions between local cortical populations of leaky integrate-and-fire (LIF) E and I point neurons receiving external input (Fig. 1a). The model receives non-local contributions from other areas. This takes the form of a depolarizing external input with a given mean value, *ν*, that is varied across simulations to set the average depolarization levels of neurons (see below). It also has an additional low frequency (largely < 10 Hz) component, whose parameters are kept constant across all simulation, which recapitulates spatially-coherent larger scale covariations across distant areas^9^. Such recurrent modeling allowed us to simulate realistic LFP and EEG proxies with characteristics such as different types of firing regimes, γ oscillations, and spectral shapes that closely resemble real cortical local networks^8,14,19,20^. From the simulated LFP or EEG data we can additionally measure key spiking outputs (i.e. firing rates) of both E and I populations.

**Fig. 1:**
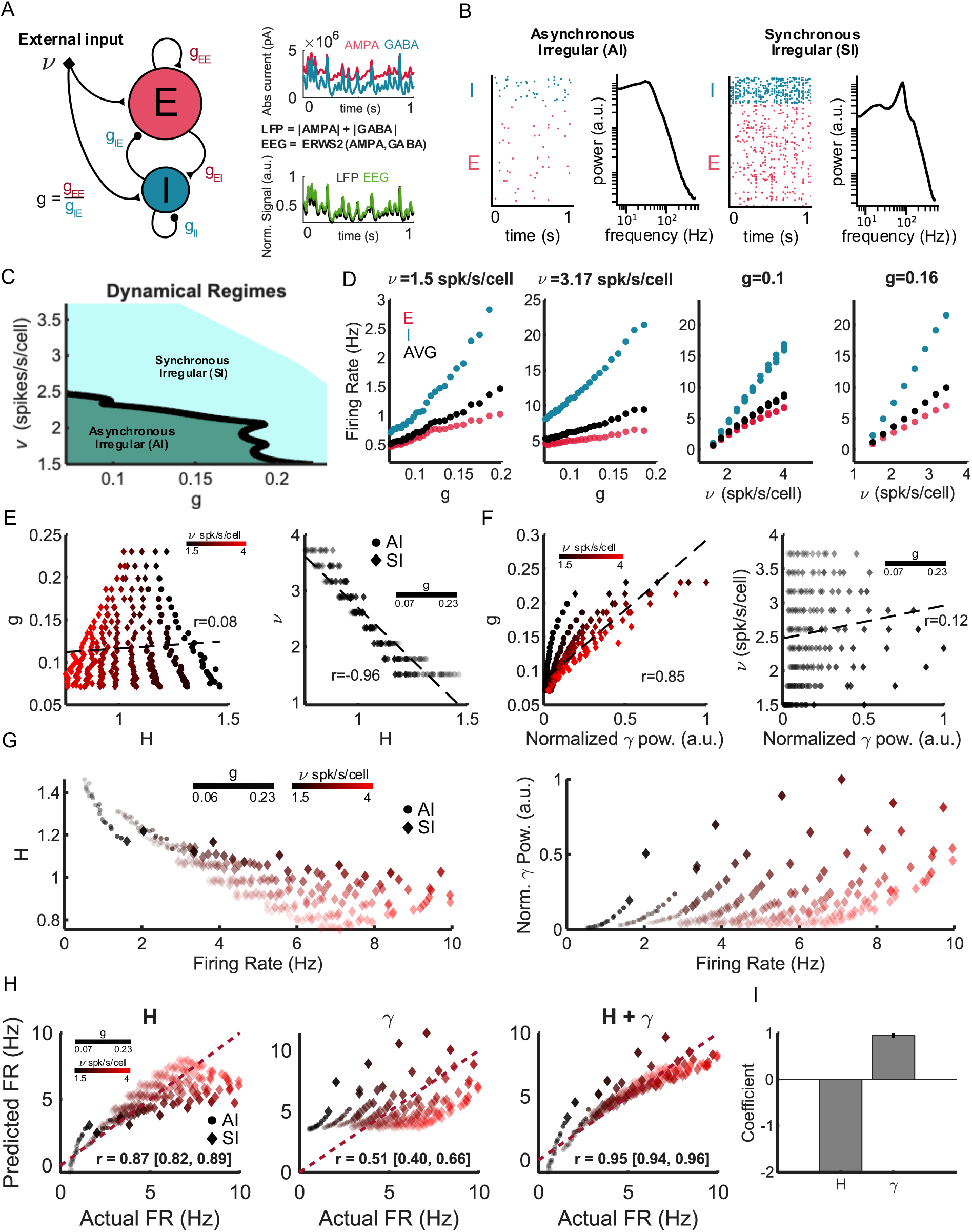
In-silico modeling showing how Hurst exponent (H) and γ power computed from LFP relate to network excitability. **a**, Schematic of the recurrently connected leaky-integrate-and-fire point neuron model used to simulate mesoscopic LFP/EEG signals while manipulating microscopic network ground truth parameters related to E and I. The model consists of 4000 excitatory (node E; red), and 1000 inhibitory (node I; blue) neurons. Neurons are recurrently connected between and within each population. We vary 2 parameters in the model: 1) the ratio (g) between excitatory AMPA (g_EE_; red) and inhibitory GABA (g_IE_; blue) synaptic conductances received by E neurons, which regulates the relative E:I interaction strength, and 2) the average input level (ν) received by both E and I neurons, which regulates the average level of excitability of individual neurons. The model was simulated over a range of g ratios that have inhibition 4-14 times greater than excitation and with average input (ν) that varied between 1.5-4 spikes per second per neuron. Examples of current traces are also shown with the different proxies used for calculating LFP (black) and EEG (light green) traces. **b,** Raster plots (left) and LFP spectra (right) of 1s of simulation time of two networks simulations which lead to asynchronous irregular (AI) and synchronous irregular (SI) firing modes. **c,** Phase diagram in the ν, g parameter space delineating SI and AI firing regions. **d,** Firing rate levels for E (red) and I (blue) neurons and for the network average (black) as a function of g and v for example parameters. **e-f,** Plots showing how H and γ vary as a function of parameters g and ν. Each dot (AI) or diamond (SI) represents the outcome of an individual simulation with the considered parameter values. The regression lines and the corresponding R^2^ are also plotted. **g,** Plot showing how the network average firing rates depend on simulation parameters. Each point represents the outcome of an individual simulation with the considered parameter values. **h,** Plot across simulations of the true actual average firing rate (x-axis) against predicted firing rates (y-axis) by a linear model using only H (left), only γ (middle) or both H and γ (right). The R^2^ (with 95% CIs computed from 5-fold cross-validation) and the diagonal line are also plotted. In panels E-H dots and diamonds represent simulations within the AI and SI firing regimes respectively. **i,** Regression coefficients of the best firing rate prediction model using both H and γ. Error bars represent 95% CIs from 5-fold cross-validation.

We independently manipulate two ground truth microscopic neural parameters that are important in driving E:I balance: 1) the ratio of excitatory AMPA (*g_EE_*) versus inhibitory GABA (*g_IE_*) synaptic conductances on E neurons (i.e. a parameter called *g*, where *g* = *g_EE_*/*g_IE_*) and 2) the average external input to the network (i.e. a parameter called *ν*). The external input *ν* parameter regulates the excitability level at which single neurons operate on average. The higher the level of *ν*, the closer the neuron’s average membrane potential will be to the firing threshold. In our simulations we vary *ν* between 1.5 to 4 spikes per second per neuron. Thus, manipulating *ν* allows us to control single neuron excitability within the network. In contrast, the *g* parameter can be thought of as a network interaction parameter as it regulates the relative strength of E-E vs I-E synapses rather than regulating the average excitability of individual neurons. In our work we vary *g* across a wide range, from 4 to 14 times greater I than E. This range covers the inhibition-dominated regime often reported in cortical data^21–24^. Overall, the combination of simulated ranges of *ν* and *g* allowed us to simulate neural activity within firing regimes characterized as ‘asynchronous irregular’ (AI; irregular Poisson-like firing alongside weak correlation between neurons) or ‘synchronous irregular’ (SI; irregular firing with moderate to strong correlations between neurons that reflect the common locking of neurons to γ oscillations)^13,19,20^ (Fig. 1b-c). Changes to both *g* and *ν* can alter the firing rate of the neurons and the same firing rate level can be achieved with smaller input *ν* and higher *g*, or with higher input *ν* and smaller *g* (Fig. 1d). Moreover, parameter changes that promote excitation by increasing either *g* or *ν* will also promote an increase of I firing because I and E are tightly coupled. This in turn means that the I firing rate will be even larger than the E firing rate increase because I neurons are less numerous than E neurons and thus need to fire more to keep up. Rather than considering the firing of E and I neurons separately, one important output defining the operating point of the network is the firing rate averaged over all E and I neurons, referred to hereafter as the average firing rate (FR). This output captures the combined effect of both changes in single neuron excitability and E:I interaction strength and will thus be taken henceforth as our key measure of ‘network excitability’.

Having discussed the characteristics of our modeling approach, we next move on to show how different components of the LFP or EEG signal depend on single neuron excitability (i.e. manipulated via *ν*) and the ratio of synaptic E versus I conductances (i.e. *g*). The results reported here are for simulated LFP data and are almost identical to those for simulated EEG proxies (Supplementary Table 1). Aperiodic or fractal components of the signal summarized by the Hurst exponent (H) (Fig. 1e; Supplementary Table 1) are highly sensitive to single neuron excitability (i.e. *ν*), but are far less predictive of the synaptic conductance ratio *g* over the entire range of simulated parameters (Fig. 1e; Supplementary Table 1). Similar effects appear for 1/f slope, albeit much less strong than for H (Supplementary Table 1). These results showing that aperiodic/fractal components are sensitive to *ν* and not *g* are surprising since they stand in contrast to prior observations using uncoupled^10^ or coupled models^11^ suggesting that aperiodic/fractal components track with *g* - that is, as *g* increases, H decreases and 1/f slope flattens. However, only fixed low *ν* input levels (i.e. *ν* = 1.5) were investigated in such prior work, making it impossible to reveal any relationship with variation in *ν*. While the current work replicates effects at *ν* = 1.5 ^10,11^, we show that this effect does not hold across all *ν* input levels. As shown in Fig. 1e and Extended Data Fig. 2c, an opposite relationship manifests when *ν* increases - that is, at higher levels of *ν*, when *g* increases, H also increases and 1/f slope steepens.

The mechanism explaining prior observations of an aperiodic/fractal-*g* relationship at low *ν* input levels (i.e. *ν* = 1.5) is very different in uncoupled^10^ versus coupled^11^ models. In Gao et al.,’s uncoupled model^10^, increasing *g* would increase the proportion of E over I synaptic currents. Because AMPA E currents simply decay faster than GABA I currents, this results in the E current contributing more to power at higher frequencies when increasing *g*, causing a flattening of the aperiodic slope of LFP/EEG. In contrast, when *g* increases within recurrently coupled networks^11^, the firing of both E and I neurons increases. However, I neurons have to increase their firing more to keep up with E neurons because they are less numerous (Fig. 1d). Thus, increasing *g* will increase recurrent I current more than recurrent E currents, which is the opposite to what Gao et al.,^10^ proposed. However, increasing *g* will still lead to a decrease in H and flattens the 1/f slope because local recurrent E and I currents contribute proportionally more to higher frequencies than to lower frequencies. This is because lower frequencies comprise additional components from large scale covariations across distant areas^9^. This means that lower spectral frequencies are proportionally less enhanced by local increases in neural firing and the associated increases of local recurrent synaptic currents. Ultimately, these phenomena asymmetrically affect higher frequencies and result in a flattened spectrum characterized by less negative 1/f slope, decreased H and faster dynamics (Extended Data Fig. 2ab-left), thus explaining why such measures correlate well with *g* under the low *ν* input regime (Extended Data Fig. 2c-left).

The ability to vary single neuron excitability via the *ν* parameter allowed us to observe other effects on electrophysiological time-series features. At high levels of single neuron excitability (i.e. *ν* = 4), neurons now must interact more strongly than at lower levels (i.e. *ν* = 1.5). In this situation, increasing *g* will notably create γ oscillations (Extended Data Fig. 2a-b). Over the entire simulated range of *ν* and *g* parameters, we find that γ power in the range of 30-50 Hz tracks well with *g* (Fig. 1f; Supplementary Table 1). This strong association with *g* is expected because γ oscillations are only generated by real instantaneous local interactions between E and I neurons. Interactions between E and I neurons are known to generate γ oscillations with a slower peak (∼20-40 Hz), while I-I interactions generate oscillations with a faster peak (∼70-80 Hz)^13^. Therefore, increasing *g* in a high *ν* context will increase E over I conductances and thereby increase more E-I than the I-I loops (Extended Data Fig. 2ab-right). This ultimately leads to relatively slower spectra with higher *g* and thus cancels or overcomes the opposite effect due to flattening of the spectra described above (Extended Data Fig. 2c-right). While γ power tracks well with *g*, we also find that it is also modulated to a much lesser extent by *ν* (Supplementary Table 1). This occurs because γ oscillations are stronger for higher *ν* input levels because synapses are more effective at higher *ν* input levels^23^.

### H and γ are complementary in predicting network excitability

Having linked H and γ to *ν* and *g* respectively, we next examined how such features predict overall ‘network excitability’, operationalized as the average FR of the network. Over our wide range of simulations, we find that H is anti-correlated with the average FR - that is, higher FR is related to decreased H (Fig. 1g-left). However, for the same reasons explained above, on a finer level, the strong anti-correlation between H and FR at fixed low input levels (i.e. *ν* = 1.5; AI regime) that are controlled by *g*, switches to weak positive correlations in contexts with high rates of input excitability (i.e. *ν* = 4; SI regime) that are also dictated by *g*. Thus, the relationship between H and overall network excitability (i.e. average FR of the network) is strongly predicted by single neuron excitability (*ν*), but is also input-level dependent when varying levels of *g*. In contrast to fractal measures such as H, periodic signal components such as γ power correlates positively with the average FR across all simulations - that is, higher γ power is always associated with higher FR (Fig. 1g-right).

H and γ are both predictive of overall network excitability, yet they are sensitive to different underlying E:I mechanisms (i.e. H-*ν*, γ-*g*). This suggests that they are complementary in their effect at predicting overall network excitability. To assess this possibility, we used a cross-validated linear regression model with actual FR as the dependent variable and H or γ as predictors. On their own, H or γ can significantly predict the FR reasonably well, but with H considerably outperforming γ (Fig. 1h; Supplementary Table 2). We next fitted a bilinear regression model using both H and γ as predictors. Here we found that using both H and γ leads to the best predictions of FR compared to models that only use H or γ in isolation (Fig. 1h; Supplementary Table 2). Furthermore, the regression coefficients show opposite signs for H and γ respectively, indicating that they contribute to predicting FR in different directions (Fig. 1i). This result suggests that both H and γ are indeed electrophysiological features of differential relevance to E:I mechanisms and provide complementary information about overall network excitability.

While we have mainly focused on features such as H and γ, we also evaluate other features, such as the 1/f slope^10^ and the total broadband spectral power^12^ to better understand if these features are better than H at predicting *ν*, *g*, and FR. Strong correlations with *ν* are observed for 1/f slope and total broadband power, but these correlations are smaller than those observed for H (Supplementary Table 1). In contrast, correlations are weak between these features and *g* (Supplementary Table 1). H, 1/f slope, and total broadband power were also strongly predictive of FR (Extended Data Fig. 2d, Supplementary Table 2). H tends to slightly outperform 1/f slope (Supplementary Table 2). However, total broadband power outperforms both H and 1/f slope (Supplementary Table 2). This result can be explained by the fact that in our model LFP power is proportional to the number of local synaptic events, which in turn increases with the FR. However, *in-vivo* total broadband power includes also significant contributions from low-frequency large-scale brain-state fluctuations. While our model does include such contributions, these are held constant during simulated variations of *ν* and *g* and therefore do not confound the above *in-silico* FR estimation. In contrast, *in-vivo* changes in local excitatory and inhibitory balance also modulate long-range interactions, producing low-frequency fluctuations that vary differently from local high-frequency activity^25^. We anticipate that this will confound FR estimation from total broadband power in real data.

In sum, we found that within simulated aggregate electrical potentials (i.e. LFP or EEG) of coupled local cortical networks, fractal components of the electrophysiological time-series (i.e. H, 1/f slope) track much better with single neuron excitability (i.e. *ν*) than with the E:I synaptic conductance ratio (i.e. *g*). In contrast, γ power tracks better with the synaptic E:I ratio (i.e. *g*) than single neuron excitability (i.e. *ν*). Overall network excitability, an important measure of E:I balance indexed by the average FR, can be best estimated using a bilinear regression of fractal (H) and periodic (γ) components of EEG/LFP spectra. However, because our modeling approach simulates local neuronal networks, it generates oscillations primarily within the γ band. Other slower spectral periodic components indicative of other non-local interactions could not be assessed. However, within the *in-vivo* experiments described in the next section, we could empirically assess whether other periodic components beyond γ-band activity could help better decode E:I balance.

### In-vivo evidence that chemogenetically altering E or I drives predicted changes to LFP H and γ

Our modeling thus far (Fig. 1) has predicted that electrophysiologically-measured H tracks with single neuron excitability, while γ tracks with the synaptic E:I conductance ratio. We next tested these predictions by examining how H and γ features behave *in-vivo* across 3 different chemogenetic DREADD manipulations in mice, whereby network excitability was altered in different ways. In all 3 experiments, we performed electrophysiological recordings. We measured multi-unit activity (MUA) within the chemogenetically manipulated area to estimate firing rates. We also measured LFPs, from which we computed H, γ, 1/f slope, and total broadband power. These 3 DREADD experiments were designed to either 1) decrease network excitability by silencing both E and I neurons (hSYN-hM4Di) or increase network excitability by 2) enhancing excitability of excitatory neurons (CamkII-hM3Dq) or through 3) inhibiting fast-spiking parvalbumin-positive (PV+) interneurons (PV-hM4Di). To help solidify predictions for these *in-vivo* experiments, we used further *in-silico* modeling to align our simulations with the physiological mechanisms of chemogenetic tools. Specifically, we manipulated bοth *g* and the resting membrane potential (via a parameter called *E_l_*) in a bespoke manner relevant to each chemogenetic experiment (Extended Data Fig. 3). This approach of manipulating *E_l_* captures the hyperpolarizing or depolarizing effects of DREADD-induced ion channel activation. These simulations showed that hSYN-hM4Di is predicted to decrease FR, increase substantially H, and produce a very small decrease in γ power (Extended Data Fig. 3a). CamkII-hM3Dq is predicted to increase FR, decrease H substantially, and produce a very small increase in γ power (Extended Data Fig. 3b). Finally, PV-hM4Di is predicted to increase FR, alongside a small increase in H, and a very large increase in γ power (Extended Data Fig. 3c).

In the first chemogenetic experiment (hSYN-hM4Di) we aimed to decrease network excitability by chemogenetically silencing both E and I neuronal populations via virally expressing DREADD hM4Di receptors in mouse medial prefrontal cortex (PFC) under the control of the hSyn promoter. This manipulation reduces excitability via DREADD-induced membrane hyperpolarization^26^, but also via a secondary dampening of synaptic release^25,26^. Thus, as expected the manipulation produced a reduction in the manipulated region of MUA FR spiking activity (Fig. 2c; Supplementary Table 3) - a measure of total local firing^8,27^. We also observed the predicted increase in H, as well as a decrease in γ power (Fig. 2c; Supplementary Table 3). Differentially-weighting H and γ in a bilinear regression model, we also were successful in predicting decreased FR levels at a group-level (Fig. 2c; Supplementary Table 3).

**Fig. 2:**
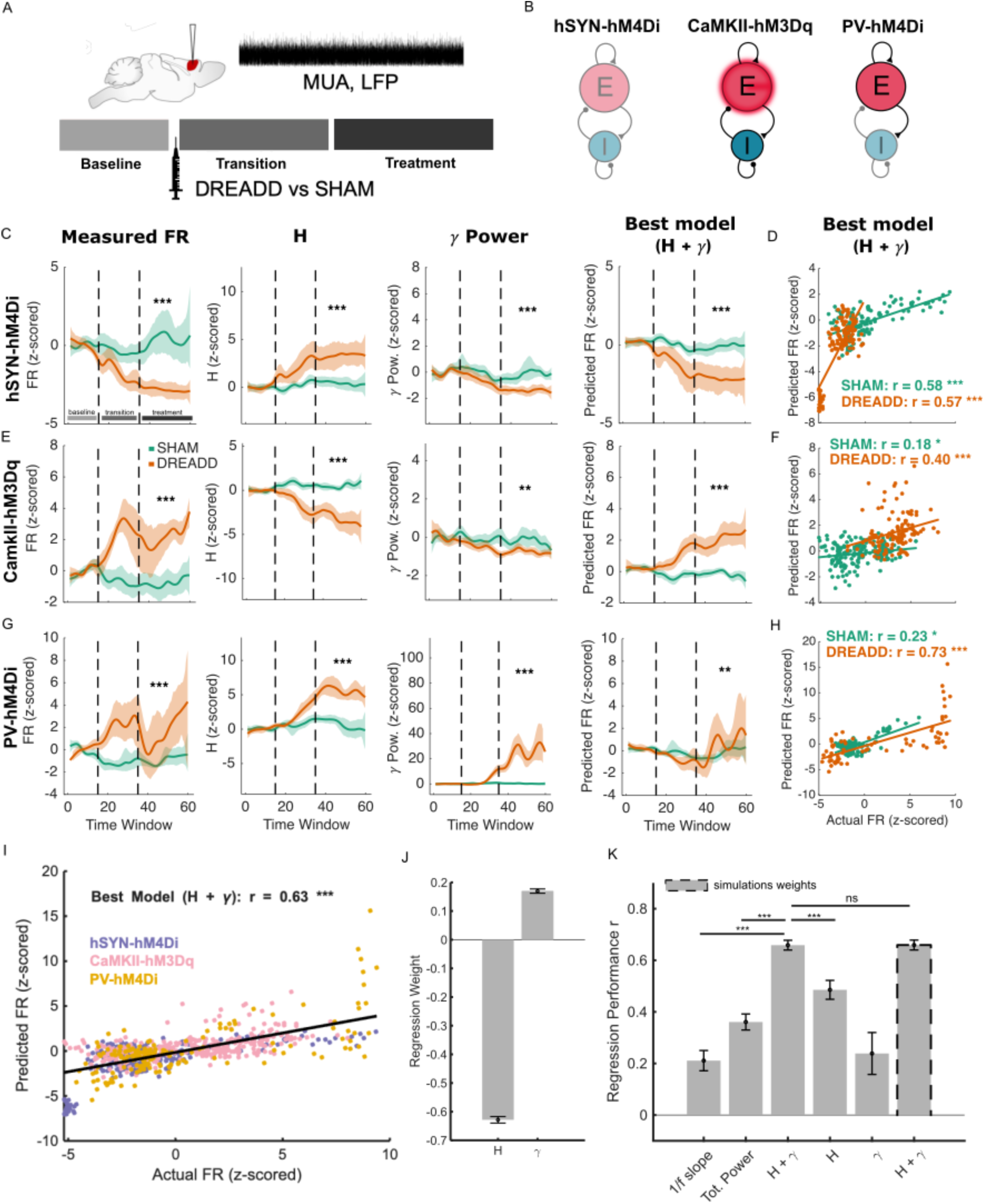
In-vivo chemogenetic manipulations that alter network excitability. **a**, Schematic of the general experimental design for DREADD experiments. The first 15 minutes of the experiment was defined as the ‘Baseline’ phase (light-gray), preceding injection of the DREADD actuator CNO to both DREADD and SHAM animals. The ‘Baseline’ period was followed by a 20-min ‘Transition’ phase (medium-gray) during which CNO is predicted to begin taking effect. After the ‘Transition’ phase is the ‘Treatment’ phase (dark-gray), which extends to 60 minutes from experiment onset and is the phase where the DREADD drug is predicted to have maximal effect. Multi-unit activity (MUA) and LFP were recorded from the injection site in PFC throughout the experiment. **b,** Schematic illustrating the effects of chemogenetic manipulations at the microcircuit level. Varying levels of transparency on E and I populations indicate the degree to which specific neuronal populations are affected: increased transparency represents stronger hyperpolarization (i.e. reduced activity), while red shading represents depolarization (i.e. increased activity). Synapses (i.e. edges within or between E and I populations) are also depicted as being affected using the same transparency coding scheme. **c,** Results for the hSYN-hM4Di DREADD silencing experiment. Trajectories of the baseline-z-scored multi-unit firing rate (far left), H (2nd to left), γ power (2nd to right) and firing rate prediction (far right) from the model using H + γ. The trajectories are computed over sliding time windows (x-axis) with each time window representing a 4 second segment of time and with each window separated by 1 minute. **d,** Scatterplot of baseline z-scored measured (x-axis) vs predicted firing rate (y-axis) from different timepoints and animals in the treatment phase of the hSYN-hM4Di experiment (DREADD,orange; SHAM, green). Insets report Pearson r between actual and predicted firing rates in each condition. **e-f,** Same as C and D but show results from the CamkII-hM3Dq experiment. **g-h,** Same as **c-d,** but for the PV-hM4Di experiment. **i,** Actual (x-axis) versus predicted (y-axis) baseline-normalized firing rates for all experiments (color-coded) and both conditions during the treatment phase of the experiment. Each dot represents a prediction for each timepoint and animal generated by the best model that utilizes both H + γ. The solid line indicates the robust fit, with the corresponding Pearson correlation coefficient (r) and p-value reported. **j,** Weights from the robust fit for the model parameters H and γ. Bars represent the mean values across the 10-fold cross-validation, and error bars indicate the standard deviation across folds. **k,** Regression performance, expressed as the Pearson correlation between predicted and actual baseline-normalized firing rates (same dataset as in panel **i**) for both DREADD and SHAM conditions during the treatment phase. Bars represent the mean correlation values, and error bars indicate the standard deviation across the 10 folds. The bar on the far right with a dotted outline represents the firing rate prediction model with weights for H and γ that are derived from in-silico modeling shown in Fig. 1i.

In a second chemogenetic experiment (CamkII-hM3Dq) we tested whether upregulating network excitability via enhancing the excitability of excitatory neurons would result in increased FR, decreased H, and increased γ (Extended Data Fig. 3b). To achieve this we chemogenetically upregulated the excitability of primarily pyramidal neurons via overexpression of hM3Dq DREADD receptor under the control of CamkII-promoter in the PFC. This DREADD manipulation produced the expected robust increase in MUA FR (Fig. 2e; Supplementary Table 3). We also observed the predicted and marked decrease in H (Fig. 2e; Supplementary Table 3), alongside a very small decrease in γ power. This slight decrease in γ power was not predicted by the model, and may be linked to the fact that CaMKIIα may have small subtle effects on a subset of other (i.e. inhibitory) cell types^28^ that we did not include in the model. Nevertheless, utilizing differentially-weighted H and γ in a bilinear FR prediction model, we successfully predicted the direction of increased FR at a group-level (Fig. 2e; Supplementary Table 3).

In a third chemogenetic experiment (PV-hM4Di), we tested whether increasing network excitability via downregulating excitability of fast-spiking inhibitory neurons would lead to the predicted increased FR, increased H, and increased γ (Extended Data Fig. 3c). Here we used a hM4Di-based chemogenetic inhibition of parvalbumin-positive (PV+) fast-spiking interneurons using Cre-dependent DREADD expression in PV::Cre mice^29^. By selectively expressing hM4Di in PV+ neurons we sought to decrease the excitability of these cells, via the combined effect of DREADD-induced hyperpolarization and reduction in synaptic release, thus weakening inhibitory synaptic transmission. Empirical electrophysiological measurements showed that inhibition of PV+ cells resulted in the predicted increased MUA FR (Fig. 2g; Supplementary Table 3). In accordance with our predictions (Extended Data Fig. 3c), this manipulation produced an increase in H (Fig. 2g; Supplementary Table 3) and also induced a major increase of γ power (Fig. 2g; Supplementary Table 3). We also found that the resulting increased *in-vivo* FR levels could be inferred by combining information about γ and H at a group-level (Fig. 2g; Supplementary Table 3).

For all three DREADD experiments, we additionally evaluated the 1/f slope and total broadband power. In the hSyn-hM4Di and CaMKII-hM3Dq experiments, both 1/f slope and total power were affected by the manipulations, though much less so than H (Extended Data Fig. 4a,c; Supplementary Table 3). The weaker effect for 1/f slope may be linked to the fact that its estimation may be more variable than that of H (Supplementary Fig. 1). In the PV-hM4Di experiment, the 1/f slope changed in the opposite direction from what would be expected (Extended Data Fig. 4e; Supplementary Table 3) given the empirically observed changes in FR (Fig. 2g), while total power was not significantly affected by the manipulation (Extended Data Fig. 4e; Supplementary Table 3). The contrast of success using total power *in-silico* versus the lack of success here *in-vivo* is likely explained by the fact that these empirical manipulations also change inter-area communication which in turn changes low-frequency components, which are instead kept fixed in our earlier modeling. Overall, the observed empirical limitations of using 1/f slope and total power suggest that H is relatively a more robust biomarker of underlying E:I-related changes. Given the success of H and γ in predicting FR, we also examined whether adding other periodic oscillatory frequency bands (e.g., δ, θ, α, β) would improve our regression models predicting FR. However, including these additional bands did not significantly increase the variance explained beyond that already captured by H and γ (Supplementary Table 5).

The comparison of SHAM vs DREADD conditions in the chemogenetic experiments shows that increases or decreases in FR can be detected by changes in LFP spectral features such as H and γ. However, it may be argued that the large variations in network configuration provided by the manipulation are unlike individual differences in naturally occurring variation across time or across animals. Thus, to evaluate whether spectral features can track individual differences on a trial-by-trial basis, we computed correlations separately for each condition between actual and predicted FR from the bilinear regression using H and γ. We found that the association between actual versus predicted FR was significant in each and every experiment and condition (Fig. 2d,f,h Supplementary Table 4) and remained significant when pooling data across all experiments (Fig. 2i; Supplementary Table 4). In contrast, models using H or γ in isolation were substantially less accurate (Fig. 2k; Supplementary Table 6-7). In sum, a bilinear estimation based on H and γ has predictive power not only for the larger group-level effects, but also for individual differences. Importantly, the regression weights were highly consistent across empirical LFP data (Fig. 2j) and simulations (Fig. 1i), with a large negative coefficient for H and a smaller positive coefficient for γ. As a result, actual FR can be accurately estimated using regression weights derived from either *in-silico* simulations or *in-vivo* recordings (Fig. 2k; Supplementary Table 6). This result adds further credibility to the *in-silico* model’s ability to capture properties of real cortical data as here we show that regression weights obtained from simulated data can be successfully applied to real data never seen before during the regression model’s training. In contrast to using H, we also considered individual differences in predicting FR using 1/f slope and total broadband power. 1/f slope significantly predicted FR in the correct direction in only 3 out of 6 cases (Extended Data Fig. 4b,d,f; Supplementary Table 4) and always underperformed H (Fig. 2k; Supplementary Tables 7). Similarly, total broadband power underperformed compared to H (Fig. 2k; Supplementary Tables 7) and significantly predicted FR in the correct direction only in the hSYN-hM4Di experiment (Extended Data Fig. 4b,d,f; Supplementary Tables 4). Together, these results indicate that H is relatively a more reliable feature for predicting FR *in-vivo* compared to 1/f slope and total power.

In sum, our experimental results support the idea that combining electrophysiologically-measured H and γ biomarkers can successfully track both increases and decreases in FR induced by chemogenetic perturbations and can also capture individual differences. In particular, combining H and γ enables accurate decoding of firing rate changes, including in regimes with large γ power shifts. Together with the *in-silico* modeling results, these findings show that measuring H and γ in LFP and EEG can reasonably predict up- and down-regulations in network excitability (i.e. FR), a key aspect of E:I balance.

### Identifying EEG-derived autism E:I neurosubtypes

Having established *in-silico* and *in-vivo* evidence that H and γ measured in LFP and EEG are driven by changes in different, yet complementary, underlying mechanisms involved in E:I balance (i.e. single neuron excitability, synaptic E:I conductance ratio), we next translationally extended this work to humans and tested whether the population of male idiopathic autism can be split into EEG-derived E:I ‘neurosubtypes’. Here we applied unsupervised stability-based relative clustering validation analysis (*reval*)^30^ to a relatively large EEG dataset of male autistic individuals (n=286) 5-21 years of age from data releases 1-10 from the Child Mind Institute Healthy Brain Network (CMI-HBN) dataset^31^. Genetic data was not available for analysis on this sample, and thus our usage of the term ‘idiopathic’ refers to the lack of knowledge regarding known genetic causes of autism. EEG data was examined under the two resting state conditions - eyes open (EO) and eyes closed (EC). H and γ were estimated across 93 scalp electrodes, averaged over 5 repeat blocks in EO and EC each, and then input into the *reval* clustering pipeline.

For both EO and EC conditions, we find that the optimal number of subtypes is 2 (Fig. 3a). Generalization accuracy for this 2-subtype solution in the held-out CMI-HBN validation set is 93% in EO and 95% in EC. These 2-cluster solutions are indicative of true clusters, as they heavily deviate from a single multivariate Gaussian null distribution (*SigClust p* < 0.05; Supplementary Table 8) and under random permutations of subtype labels (permutation *p*<0.05). Remarkable consistency was apparent in autism subtypes defined in either EO or EC conditions, with around 80% of individuals receiving the same subtype label between conditions (Fig. 3b). The obtained k=2 clustering solutions are also remarkably consistent in being identified a vast majority of time as the optimal solution over many repeat analysis runs whereby other methodological choices and parameters are varied, such as different random seeds to partition of the data into independent training and validation sets, different clustering algorithms, and different UMAP parameters (Supplementary Table 8). To further evaluate out-of-sample replicability of these autism E:I neurosubtypes, we trained a classifier on the CMI-HBN dataset EO condition and then applied it to an independent replication dataset (IIT LAND; autism n=65; typically-developing (TD) n=50) from another country/culture (Italy). The IIT LAND dataset was also collected under abstract naturalistic movie viewing conditions (‘Inscapes’) which can be a useful intermediate condition to resemble resting state conditions, while also enhancing compliance for data collection in children^32^. We find that the CMI-HBN EO classifier could identify the same 2 autism subtypes in the IIT LAND dataset with a high level of accuracy (i.e. 98%) (Fig. 3a).

**Fig. 3:**
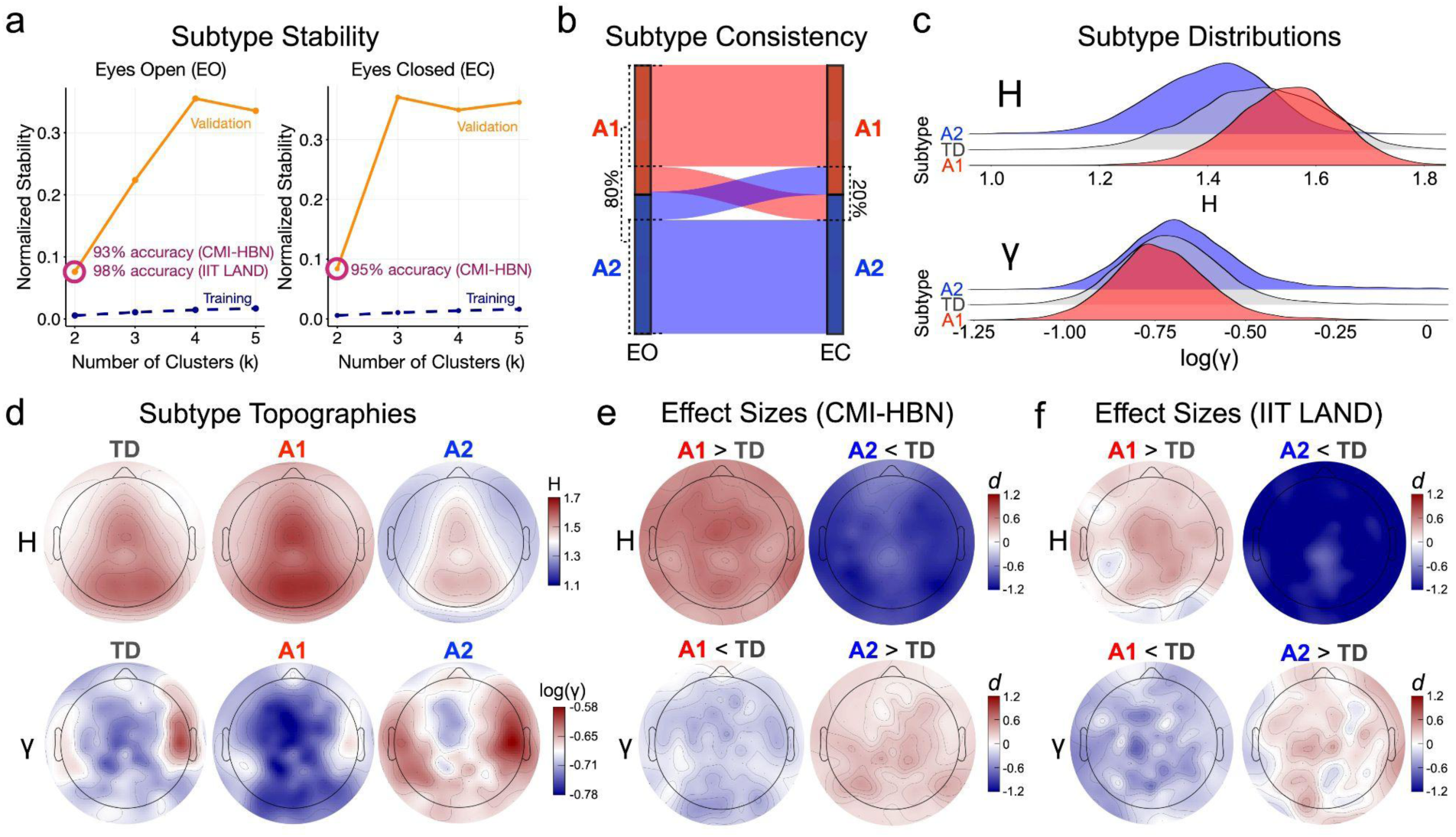
Identification of autism E:I neurosubtypes. **a**, Normalized stability plots (eyes open (EO), left; eyes closed (EC), right) indicating that a 2-cluster solution is the optimal solution that minimizes normalized stability and produces high generalizability accuracy in independent CMI-HBN validation (EO: 93%; EC: 95%) and IIT LAND replication (98%) datasets. **b,** Alluvial plot indicating how consistent A1 and A2 subtypes are across EO and EC conditions. **c,** Density plots of H (top) and γ (bottom) to describe the separation between TD (gray), A1 (red), and A2 (blue) subtypes. **d,** Scalp topographies for H (top) and γ (bottom) during EO for TD, A1, and A2 subtypes. **e,** Standardized effect size (Cohen’s d) scalp topographies for H (top) and γ (bottom) for A1 vs TD (left) and A2 vs TD (right) comparisons in the CMI-HBN dataset. **f,** Effect size scalp topographies for similar group comparisons for the IIT LAND replication dataset.

In contrast to autism-only subtyping analyses, we also attempted clustering within the CMI-HBN TD group only (n=123). Such analyses can test the null hypothesis that TD is a single population without evidence of robust and stable subtypes. While k=2 did appear as the optimal number of clusters a majority of the time, such a solution could not generalize in held-out validation data with high levels of accuracy (e.g., EO: mean accuracy over all runs = 64%) and commonly did not reject the single Gaussian null hypothesis on a majority of analysis runs (e.g., 59.6% of analysis runs with *SigClust* p>0.05) (Supplementary Table 8). These null clustering results within the TD group support retaining the underlying baseline assumption about TD as a singular population without robust and stable subtype structure in EEG H and γ features. Contrasting the lack of success in clustering TD with the remarkably robust and stable clustering in autism-only, there is strong justification for clustering within autism-only and proceeding by statistically comparing the autism subtypes to the TD group as a whole.

We next describe the autism neurosubtypes and how they deviate from TD. The A1 subtype comprises ∼47% of the autism sample and is characterized by an ‘inhibition-dominant’ profile of higher H and lower γ compared to the TD group (EO H: *Cohen’s d* = 0.58; EO γ: *Cohen’s d* = −0.24; EC H: *Cohen’s d* = 0.62; EC γ: *Cohen’s d* = −0.32; Fig. 3c-e; Extended Data Fig. 5; Supplementary Table 9 for full reporting of statistics). In contrast, the A2 subtype comprises ∼53% of the autism sample and is characterized by an ‘excitation-dominant’ profile of lower H and higher γ relative to TD (EO H: *Cohen’s d* = −0.88; EO γ: *Cohen’s d* = 0.23; EC H: *Cohen’s d* = −0.83; EC γ: *Cohen’s d* =0.33; Fig. 3c-e; Extended Data Fig. 5; Supplementary Table 9). The opposing directionality of differences between H and γ is important to underscore as these opposing directions are similar to the trade-off between H and γ observed *in-silico* and *in-vivo* models (Fig. 1i, 2k). This trade-off between H and γ observed in the A1 and A2 subtypes in CMI-HBN dataset was also identified in the A1 and A2 subtypes identified in the IIT LAND replication dataset (Fig. 3f), with a vast majority of all EEG channels showing congruency of effect size directionality between CMI-HBN and IIT LAND datasets (Fig. 3f; Extended Data Fig. 6).

### Subtype differential brain-behavior relationships

We next sought to test for phenotypic differences between these autism E:I neurosubtypes. Amongst n=138 phenotypic variables over 13 phenotypic domains (see Supplementary Table 10 more details) we found that a majority of these variables (∼64%) showed evidence of strong case-control differences, but without remarkable differences between A1 and A2 subtypes (Extended Data Fig. 7). However, there were some subtle subtype-specific differences to highlight. A2 had a tendency of showing lower performance on some cognitive measures of attentional and inhibitory control, processing speed, visual spatial skills, and fluid reasoning. In contrast, A1 had specific issues compared to TD within some variables across demographic, family, motor, physical, comorbidity, sleep, and addiction domains (Extended Data Fig. 7). Thus, although there were some subtle subtype-specific differences, most of the on-average behavioral differences were similar in autism subtypes.

In addition to testing for on-average differences in phenotypic measures, we also assessed brain-behavioral relationships with partial least squares (PLS) analysis. PLS allowed for testing relationships between multivariate neural (i.e. H or γ) and behavioral measures (i.e. n=139 variables over 13 domains). We identified one latent variable (LV1) pair in H and γ analyses respectively that were highly statistically significant after FDR correction (Fig. 4a; Supplementary Table 11). Brain salience maps were topologically very similar between EO and EC and highlight important contributions from most EEG channels (Fig. 4b-c). Within each group there was remarkable similarity in behavioral saliences across EO and EC analyses (i.e. r>0.58, p<0.05 for all diagonal values of matrices shown in Fig. 4d-e). This high level of within-group similarity across EO and EC suggests that brain-behavioral relationships are robust irrespective of resting state viewing contexts (i.e. EO, EC). In contrast to this within-group consistency, there was a striking lack of similarity in behavioral saliences for nearly all between-group comparisons (i.e. p>0.05 for all off-diagonal values of matrices in Fig. 4d-e). This result highlights a key conceptual point that brain-behavior relationships are not similar between groups. A further illustration of these between-group differences in brain-behavioral relationships can be seen in the word clouds depicted in Fig. 4f-g. These word clouds are based on the most important behavioral variables contributing to LV1 with a non-zero relationship and can be used to visualize the phenotypic domains where groups show pronounced differences in brain-behavioral relationships. Here we see that autism and many other neuropsychiatric and mental health-related domains known to highly co-occur with the autism diagnosis (e.g., mood, anxiety, ADHD, comorbidity) are either uniquely important to A1 (bolded terms in the middle column for Fig. 4f-g) or are differentially important for A1 compared to A2 (thick purple lines between middle and right columns in Fig. 4f-g). In contrast, language and cognition domains appear as uniquely important for A2 (Fig. 4f-g). We also see that physical and demographic domains tend to show opposite directionality of brain-behavioral relationships between TD and A1 across both H and γ (Fig. 4f-g).

**Fig. 4:**
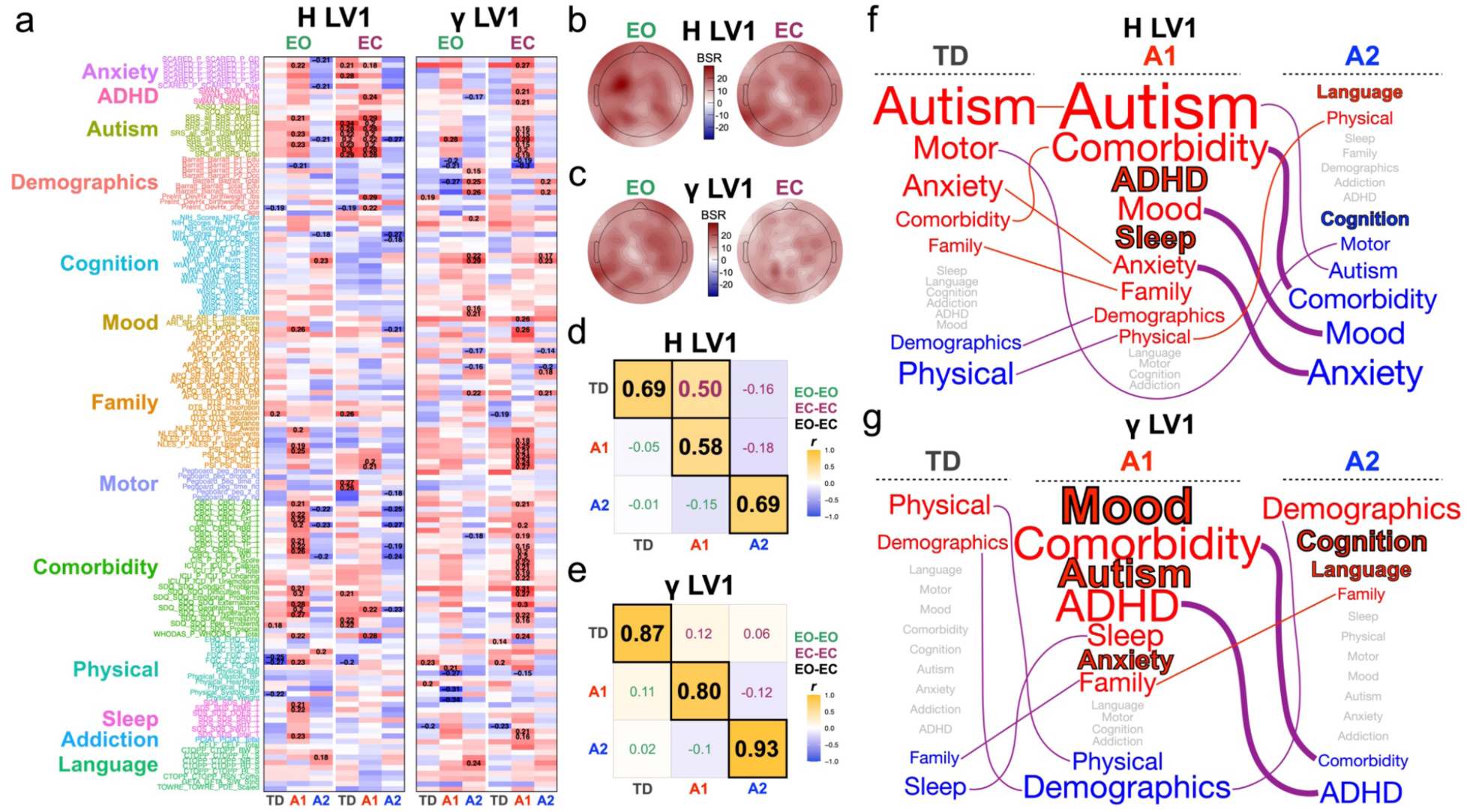
Subtype differential brain-behavior relationships. **a**, Heatmaps of behavioral salience matrices for the LV1 result from 4 PLS analyses whereby H or γ were the neural features measured under EO or EC contexts. The rows of these matrices show each behavioral variable, and the coloring of the labels corresponds to the 13 behavioral domain categories labeled to the left of the heatmaps. The columns of the heatmaps correspond to TD, A1, and A2 subtypes respectively. The colors in the heatmap show the correlation values for each variable and cells with the number indicate variables of most importance contributing to the LV1 relationship (i.e. non-zero variables) whereby their 95% bootstrap confidence intervals do not encompass a null value of 0. **b-c,** Brain bootstrap ratio (BSR) scalp topographies (EO, left; EC, right) for H LV1 (b) and γ LV1 (c) respectively. BSR values indicate the importance of EEG channels in the overall LV1 relationship, with values above 2 indicating significant importance. In these plots all channels have BSRs well over 2, indicating that all channels are significantly important to the relationship. **d-e,** Correlation matrices quantifying the degree of similarity between behavioral saliences within-and between-groups. Panel **d** shows the similarities for H LV1, while panel **e** shows the similarities for γ LV1. The diagonal values (black font color) indicate within-group but between-condition (EO-EC) similarities. The lower triangle (green font color) represents between-group similarity within EO (EO-EO), while the upper triangle (purple font color) represents between-group similarity within the EC (EC-EC) condition. Statistically significant correlations are emphasized in bold and with larger size and by black rectangles around the cell. **f-g,** Word clouds (panel **f** for H LV1; panel **g** for γ LV1) to emphasize the specific non-zero relationships in panel **a** over the 13 behavioral domain categories. Red or blue text color represents the directionality of the average correlations in panel **a**. Words colored in gray represent behavioral domain categories with no non-zero relationships of importance. The size of the words reflect the weighted average of correlation in panel **a**, where the weight indicates the percentage of variables in a particular category that have a non-zero relationship. The domains in bold with black outline are domains that are unique to one of the groups. Lines between groups indicate domains that appear for more than one group, with purple indicating different directionality, whereas red and blue lines indicate the same directionality. Thick lines indicate different directionality of relationships between A1 and A2 for specific domains that are relevant for other co-occurring neuropsychiatric or mental health conditions.

## Discussion

The overarching goals of this work were twofold. First, we used extensive *in-silico* simulations alongside 3 different *in-vivo* manipulations of excitation and inhibition to investigate how specific E:I mechanisms can be decoded from different electrophysiological biomarkers measured in LFP or EEG data. The second goal was to show an example of the potential translational impact of decoding E:I mechanisms from EEG data in male autistic children. Such a goal is positioned to help gain further clarity and advance our understanding of a long-standing theory in autism research - the E:I imbalance theory^1,3^. Overall, the combined insights from the first goal using *in-silico* and *in-vivo* experiments helps to provide a novel level of mechanistic understanding that neuroscientists could use to help decode E:I mechanisms from non-invasively measured electrophysiological data. The insights gleaned from this first goal could potentially be far reaching in allowing for a large range of research using non-invasive electrophysiology in humans to examine new aspects of brain function and how they may relate to the intricate balance between E and I neuronal populations. As exemplified in our human work on autism, decoding E:I relevant mechanisms from non-invasive electrophysiological data may also have far reaching implications when applied translationally to patient populations.

In terms of mechanistic insights, this work disentangles two important E:I-relevant mechanisms, single neuron excitability and E:I interaction strength, and relates them to distinctive electrophysiological time-series features that can be measured *in-vivo* with EEG data. Our *in-silico* simulations show that fractal components of the electrophysiological time-series data measured by the Hurst exponent (H) track well with a parameter that controls average single neuron excitability, operationalized as the level of external input to individual neurons (i.e. *ν*). This single neuron excitability parameter (*ν*) regulates a neuron’s average membrane potential, with higher levels raising the membrane potential closer to the neuron’s firing threshold. In contrast, periodic γ oscillations in electrophysiological time-series data track well with the average E:I interaction strength, operationalized as the ratio of E vs I synaptic conductances (i.e. *g*). These new insights relating *ν*-to-H and *g-*to-γ stand in contrast to prior influential work that suggested that fractal components of electrophysiological data (e.g., H, 1/f slope) were primarily linked to the synaptic *g* ratio^10,11^ - that is, flatter 1/f slope and decreasing H with increasing *g*. While we found some supportive evidence of a similar relationship between H (and 1/f slope) and *g*, this relationship is restricted to contexts with low levels of single neuron excitability, when the firing rate levels are very low and the network is in an asynchronous irregular firing regime (i.e. black points in the left plot of Fig. 1g). By extending our simulations to the range of values for *ν* that go from 1.5 to 4 spikes per second per neuron, we are now able to show that this relationship between H and *g* first attenuates and then flips directionality as single neuron excitability approaches higher levels (e.g., v = 4 spikes per second per neuron). These new insights are compatible with other recent papers that have called into question a simple relationship between aperiodic features (e.g., 1/f slope) and *g*^33,34^. However, our work clarifies how we can change our interpretations about measures like H and/or 1/f slope. Rather than tracking E:I mechanisms like *g*, we show that this relationship is much more complex and interacts with levels of *ν*. Therefore, the large number of papers that claim aperiodic measures track with synaptic E:I ratio must take in this new evidence and use what we view as a more correct interpretation - that is, such measures actually track better with single neuron excitability rather than E:I interaction strength. Any such relationships between these measures (i.e. H or 1/f slope) and *g* must be contextualized to the degree of single neuron excitability in specific situations (e.g., rest, task, etc).

In contrast to aperiodic/fractal components being sensitive to single neuron excitability, the synaptic *g* ratio appears to be most sensitive to changes in periodic γ oscillations. Across the entire simulated range of *ν*, the relationship between γ and *g* always remains positive (Fig. 1g) - that is, as the synaptic E:I ratio becomes more excitation-dominant, more γ oscillations emerge. The strongest empirical example of this comes from the chemogenetic experiment whereby we increased network excitability via silencing fast-spiking parvalbumin (PV) inhibitory neurons. Without this pronounced ‘neural brake’ from PV inhibitory neurons, the strongest electrophysiological feature that changes is the pronounced increase in γ oscillatory power (Fig. 2c). In contrast, the other route to increasing network excitability was via the *in-vivo* CamkII-hM3Dq experiment to enhance excitability via effects primarily on pyramidal neurons. Increasing network excitability in this experiment did not similarly induce increased γ. Instead, γ was slightly reduced in the CamkII-hM3Dq experiment. This slight reduction in γ may be explained by subtle effects that CaMKIIα may have on a small subset of inhibitory cell types^28^. Nevertheless, the combined inferences from the PV-hM4Di and CamkII-hM3Dq experiments affirm well-known findings that γ oscillations are primarily controlled by inhibitory neurons^15^, rather than manipulations that primarily affect excitatory neurons.

Although one way to interpret the work is to focus on *ν* and *g* parameters and the EEG measures that may help decode them (i.e. H and γ), it is important to also consider the idea of ‘network excitability’ (i.e. average FR of the network) as an emergent concept that measures the operating point of the E:I network and captures both the contributions of single neuron excitability and E:I interaction strength mechanisms. As shown in Fig. 1d, both *g* and *ν* can alter the average FR of the network. Similar FR levels occur with smaller input *ν* and higher *g*, or with higher input *ν* and smaller *g*. Thus, one important finding of this work is that both H and γ are complementary and important independent contributors to help explain network excitability. The influence of H and γ on network excitability are opposites, with decreasing H and increasing γ predicting higher network excitability. These *in-silico* predictions of opposite influences of H and γ for predicting FR were further verified in the *in-vivo* chemogenetic manipulation experiments. Strikingly, FR from the *in-vivo* experiments could be equally well predicted by H and γ coefficients measured in real electrophysiological data or from coefficients from the *in-silico* simulated data (Fig. 2k). A further higher-level of empirical validation for the opposing effects of H and γ can be seen in opposing directionalities of H and γ in the autism neurosubtypes - A1 was characterized by increased H and decreased γ, while A2 was characterized by decreased H and increased γ. Thus, there is continuity across *in-silico*, *in-vivo*, and human work to showcase that different underlying E:I mechanisms contribute to the idea of E:I balance and the manifestation of such (im)balance in autistic subtypes.

Another important nuance of this work to highlight is the fact that H and γ tend to be the most powerful features that help predict network excitability (e.g., average FR). Within the *in-vivo* chemogenetic experiments, 1/f slope and total broadband power were not as successful in predicting average FR compared to H. Furthermore, when other periodic oscillatory bands are added to regression models to predict FR, they do not add to explain significantly more variance over and above H and γ (Supplementary Table 5). While such negative results may imply these other EEG biomarkers may not predict local FR, this leaves open the possibility that these other types of features may play a role in other aspects of E:I balance that are not the current focus of this investigation.

From this work, we were able to translationally utilize such EEG E:I biomarkers (i.e. H and γ) to derive data-driven autism neurosubtypes that confirm that E:I imbalance in idiopathic male autistic children spans opposing poles of E:I balance^2^ - from a relatively ‘inhibition-dominant’ (A1) to a relatively ‘excitation-dominant’ (A2) subtype. Such insights mirror opposing poles of E:I imbalance observed in highly penetrant rare genomic causes of autism^2^. However, a new insight of this work is that we can now move beyond inferences from rare genomic causes and animal models of autism and show that similar opposing poles of E:I imbalance extend to the much larger male idiopathic autism population. Because of the unlock of being able to make these characterizations in non-invasively measured EEG data, this work showcases a proof-of-concept for how the E:I theory of autism could be studied more extensively over the entire range of autistic cases and not just those with specific highly-penetrant genomic causes. Several high-impact clinical uses of such work in humans might be envisioned, including evaluation of clinical trials of pharmacological interventions targeting specific kinds of E:I mechanisms. For example, pre-treatment EEG measures could be utilized in clinical trials to identify the subset of patients that might be most affected by the specific kind of pharmacological manipulation to either E or I mechanisms. Another application of this work could be to better understand other heterogeneous phenomena that characterize multiscale functional brain organization, such as connectomics work in autism. Existing work suggests there could be a possible link between E:I balance and how functional connectivity manifests (e.g., ^25^). Examining how differing poles of E:I balance in autistic patients may help explain heterogeneous patterns of hyper-versus hypoconnectivity that is observed in different types of autistic patients (e.g., ^35^).

We also showed that autism E:I neurosubtypes are quite robustly identified after many methodological variations and within multiple validation sets, including a distal replication dataset collected in another country and with a different paradigm for measuring intrinsic resting state organization. The high level of generalizability accuracy of the subtypes within validations sets in CMI-HBN and IIT LAND suggest that these E:I neurosubtypes are a population-level stratification that is evident in idiopathic male autistic children. Our focus on males was due to limitations in sample sizes for autistic females and due to literature suggesting that E:I imbalance may be moderated by sex/gender^11^. However, future work should test the level of generalization of these subtypes in autistic females. Future work should also examine how well these results might generalize to autistic individuals at younger ages and with lower levels of language and cognitive function. While the lower bound age for the CMI-HBN dataset was 5 years, the IIT LAND dataset included individuals as young as 18 months.

While autistic males can be robustly split into these E:I neurosubtypes, we also observed that TD children cannot be robustly split into such subtypes. This null evidence supports the general assumption that the TD population is best characterized as a single Gaussian distribution with important individual differences parameterized as a spectrum of continuous variation. While some accounts of autism tend to characterize individual differences as a ‘spectrum’, this work clearly points to a first-level characterization of autism as several ‘autisms’^36^ rather than one continuous spectrum of E:I balance. Further evidence in support of the idea of multiple ‘autisms’ comes from results showing a lack of similarity in brain-behavioral relationships between autism E:I neurosubtypes. Such evidence runs counter to the idea that brain-behavioral relationships should be spectrum-like across the entire autism population. The brain-behavioral relationships also highlighted domains of importance for specific subtypes, such as language and cognition for A2, and mood, sleep, ADHD, anxiety, other comorbidities, and autism for A1. These results may be important given other work showing that subtypes differentiated by similar phenotypic domains have different genetic underpinnings that affect different E or I cell types^37^. Furthermore, autisms that are diagnosed early versus late in development tend to have differences at the genetic level linked to language and cognitive or other mental health issues^38^. The results also align well with other theoretical models about heterogeneity in autism that distinguish subtypes by early developing language and intellectual abilities^36^.

In summary, we have shown that non-invasive electrophysiological data can be utilized to decode specific E:I mechanisms. Fractal components of the signal measured by H alongside periodic γ oscillations pick up on different aspects of E:I balance. H tracks with single neuron excitability, while γ tracks with E:I interaction strength. We also show an application of these E:I-relevant EEG biomarkers applied translationally to a long-standing theory about E:I imbalance in autism. We find robust and replicable evidence that within the population of idiopathic autistic males there are two opposing E:I neurosubtypes - one that is relatively more ‘inhibition-dominant’, while the other is relatively more ‘excitation-dominant’. Further validation of these neurosubtypes and their behavioral relevance could be seen in subtle behavioral differences in A1 versus A2, but also by the stark lack of similarity in brain-behavioral relationships, despite very strong brain-behavioral relationships within-group across different resting state viewing conditions. Overall, such work has numerous potential applications for furthering work on how E:I balance relates to normative and atypical brain function, and in particular, within autism research.

## Methods

### CMI-HBN dataset

To identify autism E:I neurosubtypes, we utilized EEG and phenotypic data from data releases 1-10 of the Child Mind Institute Healthy Brain Network (CMI-HBN) dataset. The CMI-HBN is an initiative to create a biobank of individuals aged 5-21 from the New York area. Participants undergo diagnostic and phenotypic assessments, EEG and MRI scanning during resting state, naturalistic viewing, and task conditions. Data was acquired at four sites in the New York area - Staten Island, Midtown, Harlem, and a mobile unit. The CMI-HBN initiative received ethical approval by the Chesapeake Institutional Review Board, and written informed consent and assent was obtained from all participants and/or their legal guardians. For further details on the CMI-HBN initiative see Alexander et al.,^31^.

For this work we started by isolating all participants from data releases 1-10 that had a diagnosis of autism (n = 412) or who were typically-developing (TD), as indicated by the label ‘No Diagnosis Given’ (n = 257). From this pool of data, we excluded participants with missing resting state EEG data (n = 15) or whose data could not be successfully preprocessed (n = 106). Given a low sample size of female autistic participants (n = 50), female participants (n = 139) were also excluded. The final sample size was of n = 286 autistic males and n = 123 typically developing males. See Supplementary Table 12 for a summary of participant characteristics. Phenotypic behavioral data from the CMI-HBN dataset comprised n=138 variables spanning 13 different domains from anxiety, ADHD, autism, demographics, cognition, mood, family, motor, comorbidity physical, sleep, addition, and language measures. See Supplementary Table 10 for a full description of all behavioral variables.

### IIT LAND replication dataset

To test whether autism neurosubtypes identified in the CMI-HBN dataset are truly indicative of generalizable subtypes in the autism population, we examined EEG data from a completely independent dataset in Italy (IIT LAND) of autistic (n = 65, mean age = 10.99 years, SD age = 5.24 years) and TD males (n = 50, mean age = 9.82 years, SD age = 5.72 years) in the age range of 1.58-34.75 years. Participants were recruited primarily from public health service diagnostic services in the Trentino region of Italy (APSS), private centers in Trentino that offer diagnostic and intervention-related services (ODF), or via social media. This dataset was collected under ethical approval from the Comitato Etico per le sperimentazioni cliniche dell’Azienda Provinciale per Servizi Sanitari della Provincia Autonoma di Trento. All children gave assent and parents provided written informed consent according to the Declaration of Helsinki. Behaviorally, IIT LAND did not sample a wide range of phenotypic measures and thus comparisons of subtypes on similar behavioral measures as the CMI-HBN were not possible. See Supplementary Table 12 for a summary of participant characteristics.

### EEG data acquisition

EEG data in both CMI-HBN and IIT LAND datasets were recorded using a 128-channel EGI HydroCel Geodesic system (Electrical Geodesics Inc; EGI). Data was recorded at a sampling rate of 500 Hz, with a bandpass filter of 0.1 to 100 Hz and reference at the vertex of the head (Cz). Resting state data in CMI-HBN was collected for 5 minutes, while the participant sat in front of a screen with a fixation cross in the center. Throughout the recording, participants in the CMI-HBN dataset alternate having their eyes open (EO; 20 seconds blocks) and closed (EC; 30 seconds blocks) for five blocks. Resting state data collection in IIT LAND was collected using the ‘Inscapes’ paradigm^32^. Past work on Inscapes shows that it can be a substitute for measuring intrinsic functional brain organization in a manner similar to resting state^32^, while also optimizing for compliance in testing in young children. The Inscapes paradigm lasted 7 mins in total. From this data, we cut out the first 2 minutes of the preprocessed data for use in data analyses. In order to be compatible with the length of epochs measured in CMI-HBN, Inscapes data was epoched into 5 segments lasting 20 seconds each and separated by a 4.5 second gap.

### EEG data preprocessing

In order to reproducibly handle the large amount of EEG data, we developed a semi-automated preprocessing pipeline that combines a set of custom MATLAB scripts executable within the EEGLAB^39^ framework with other associated software (ASR^40,41^ and IClabel^42^). The pipeline consists of a sequence of steps. Step one consists of removal of the outer ring of channels (26 channels) that are mainly located on the face and the neck. Data was then downsampled to 250 Hz and a basic FIR filter (*filtfilt.m* function) with a high pass cutoff of 1 Hz and a low pass cutoff of 80 Hz was applied. Line noise was removed via a notch filter centered at 60 Hz +/-2 Hz for the CMI-HBN dataset and 50 Hz +/-2 Hz for the IIT LAND dataset. Bad channels were identified based on a combination of three different parameters: 1) flatline duration (for more than 5 consecutive seconds); 2) standard deviations (> 4) as compared to the total channel signal; 3) correlation with the neighboring channels (lower than the default value of 0.8). Channels falling into one of these conditions were rejected and interpolated with a spherical method. In a further step the Artifact Subspace Reconstruction (ASR) algorithm was applied to detect data segments contaminated by non-stereotypical artifacts. A preliminary calibration of the rejection threshold criteria is computed on a ‘cleaner’ portion of the EEG signal. Then ASR identifies the artifact subspace and repairs the artifactual samples based on the predefined threshold values. After re-referencing to the average of all channels, the number of channels was further reduced by 10% to match the number of available samples input into independent component analysis (ICA; with ica_type set to ‘runica’). The resulting components were classified by the ICLabel algorithm as deriving from ‘brain’ or a variety of non-brain sources (e.g., muscle, line noise, channel noise, eye, or other). The non-brain ICs (probability < 20% of being ‘brain’) were then projected out from the data. After preprocessing was complete, we output preprocessing reports that could be manually checked and classified for exclusion or inclusion into further downstream data analysis. For this data quality control step, all preprocessing reports were manually inspected by MVL and a subset of overlapping subjects were also inspected by NB and AV. From this data quality control check we excluded participants for a variety of qualitative and quantitative reasons, including issues with the raw time-series data, large numbers of bad channels or bad samples that could not be repaired, too few high probability brain independent components (IC) in the top-35 ranked ICs, and/or odd looking IC topographies amongst ICs labeled as ‘brain’. For examples of excluded and included participants, see Supplementary Fig. 2. Data quality metrics such as number of bad channels and percentage of bad samples were computed for all participants retained for further downstream analysis and were used for further denoising steps before the subtyping analysis.

### EEG subtyping analysis

For EEG-based neural features relevant to E:I imbalance, we utilized the Hurst exponent (H) and peak amplitude of gamma band (γ) oscillations. H was computed with the *bfn_mfin_ml.m* function from the *nonfractal* MATLAB library (https://github.com/wonsang/nonfractal). For γ oscillations, we used the fitting-oscillations-and-one-over-f (FOOOF) algorithm implemented in Python^6^ (https://fooof-tools.github.io/fooof/). Due to different line noise frequencies in the USA versus Europe, peak γ power was measured within the band of 30-50 Hz for CMI-HBN and 30-45 Hz for IIT LAND. Η and γ features were then averaged over blocks within EO, EC, or Inscapes conditions. After obtaining block-averaged H and γ for each participant and electrode, we implemented a final denoising procedure to remove variance associated with data quality metrics from the preprocessing such as number of bad channels and percentage of bad samples. This denoising step was implemented via a linear model with H or γ as the dependent variable and diagnosis, age, study site, number of bad channels, and percentage of bad samples as independent variables. Beta coefficients for number of bad channels and percentage of bad samples were isolated and then projected out of the data to adjust H or γ values for these metrics. This procedure was done to ensure that any residual effects of data quality were handled before subtyping analysis was implemented. A final matrix for use in subtyping analysis was made with subjects along the rows and concatenated block-averaged H and γ features along the columns.

To implement EEG-based neurosubtype stratification in autism, we utilized an unsupervised data-driven clustering approach called *reval*^30^, that seeks to identify the optimal number of clusters with optimal level of generalizability in new data. This approach is based on stability-based relative clustering validation^43^ and is explained extensively in our past work^30^. Here we will briefly reiterate what *reval* attempts to achieve as well as how it goes about identifying a solution to the problem. The *reval* approach transforms unsupervised learning into a classification problem that allows for immediate translation of data-driven clustering solutions into supervised classification models that can be empirically demonstrated to be robust and generalizable partitions of a population. The *reval* algorithm can be described in the following series of steps. First, the dataset is split into independent training (dataset ***X_tr_***) and validation (dataset ***X_val_***) sets. Within the training set (***X_tr_***), *reval* splits the data into an internal training (***X_tr_tr_***) and test (***X_tr_ts_***) set, with the goal to identify the optimal number of clusters (***k***). Clustering algorithm ***Α_k_*** is then fit to both ***X_tr_tr_*** and ***X_tr_ts_*** and produces clustering labels ***Y_tr_tr_*** and ***Y _tr_ts_*** respectively. A classifier (***Φ_tr_tr_***) is then trained on [***X_tr_tr_***, ***Y_tr_tr_***] and then fit to ***X_tr_ts_*** to predict labels ***Y_tr_ts_pred_***. Misclassification error is computed by comparing ***Α_k_***’s clustering labels (***Y_tr_ts_***) to a classifier’s (***Φ_tr_tr_***) predicted labels (***Y_tr_ts_pred_***). The *reval* algorithm iterates this procedure over a range of possible ***k*** clusters, and then identifies the optimal ***k*** that minimizes misclassification error on the internal test set (***X_tr_ts_***). Misclassification error is the normalized Hamming distance between the actual clustering labels (***Y_tr_ts_***) versus the classifier’s (***Φ_tr_tr_***) predicted labels (***Y_tr_ts_pred_***). However, because of the non-uniqueness of clustering labels, *reval* permutes the labels (***Y_tr_ts_***) and uses the Kuhn-Munkres algorithm to identify the labeling solution that minimizes misclassification error. This final measure of misclassification error is called ‘clustering stability’^43^ and this index ranges from 0-1, with lower values indicating more stable and reproducible clustering solutions. Stability is then normalized to the stability of random labelings to arrive at the final measure of normalized stability. Thus, the overall first goal in *reval* is to identify the clustering solution ***Α_k_*** that minimizes normalized stability. Once the optimal ***k*** is identified from the training set, we then train a classifier (***Φ_tr_***) on the entire training set at the optimal ***k***, and then apply it to held-out validation set (***X_val_***). Clustering labels (***Y_val_***) obtained on the validation set ***X_val_*** are then compared to the classifier’s (***Φ_tr_***) predicted labels (***Y_val_pred_***) and we compute a measure of generalizability accuracy operationalized as 1-misclassification error. Clustering solutions with high generalizability accuracy are the solutions we seek, as they are informative to solutions that have high potential to generalize well on the larger population.

Before entering the *reval* clustering analysis, the data is split into independent training (***X_tr_***) and validation (***X_val_***) sets (training set = 50%; validation set = 50%) while balancing the splits for variables such as age and comorbid ADHD diagnosis. Three preprocessing steps are then applied. First, we impute missing values with a k-Nearest Neighbors imputation algorithm where the number of neighbors is identified via grid search (sklearn.impute.KNNImputer). The next two steps are ones where the parameters are fit on the training set and then applied to the validation set - scaling each feature in the dataset to mean of 0, standard deviation (SD) of 1 (sklearn.preprocessing.StandardScaler) and applying the dimensionality reduction technique Uniform Manifold Approximation and Projection (UMAP)^44^ (https://umap-learn.readthedocs.io/en/latest; n_neighbors = 30, min_dist = 0.0, n_components = 2, metric = Euclidean) to reduce the number of features from 186 to 2. The clustering algorithm used for this work was k-means clustering and was chosen on the basis of being one of the simplest and easier to understand clustering algorithms to use and because it is the same algorithm used in further validation analyses we run such as *SigClust*^45^. The classification model used throughout *reval* for this work was chosen to be k-nearest neighbors (kNN). This type of classifier was selected based on prior evidence that it typically performs best in combination with k-means clustering in several validation analyses across multiple types of datasets, as shown in the original *reval* paper^30^.

While the *reval* clustering approach is meant to ensure the stability of clustering solutions, the optimal k that is determined by *reval* must still be tested for whether it is a solution indicative of a situation where actual true clusters exist. To test the solution against the null hypothesis that no clusters are apparent in the data, we utilized a Monte Carlo simulation framework called *SigClust*^45^ meant to test whether the data significantly deviate from the null hypothesis that the data originates from a single multivariate Gaussian null distribution. This test was implemented with the *sigclust* library in R^45^ with the number of simulations set to 10,000. On each iteration of the simulation, *SigClust* generates a single multivariate Gaussian null distribution and applies k-means clustering at a specific optimal k. *SigClust* then estimates a test statistic (CI), operationalized as the within-class sum of squares about the mean, divided by the total sum of squares relative to the overall mean. Doing this across all iterations allows for building a null distribution of the CI test statistic assuming a single Gaussian distribution. We then compare the test statistic in the real data to the null distribution to calculate a p-value that allows us to test whether the clustering test statistic significantly deviates from the null hypothesis that the data derive from a single multivariate Gaussian distribution. Additionally, we also compute this same clustering test statistic after randomly shuffling the subtype labels (e.g., 10,000 times) to compute a permutation p-value. This permutation p-value is cited in addition to the p-value from the Monte Carlo analysis conducted by *SigClust* and allows for a further test of clustering against a null hypothesis where the subtype labels are randomized.

We also undertook further analyses to evaluate the robustness of *reval* clustering results over a range of different methodological choices and subject groupings. For the methodological changes, we re-ran *reval* over 20 different random seed states for randomly partitioning the data into training and validation sets. We also re-ran *reval* over 2 types of clustering algorithms (i.e. k-means, agglomerative hierarchical clustering) and with either local or global UMAP nearest neighbor parameters (e.g., UMAPl n_neighbors = 5; UMAPg n_neighbors = 30). Results were considered robust over the 20 repeat runs of different seed states if the optimal k solution appeared for a majority of runs and in more than half of all analysis runs and showed generalizability accuracy >80%. Furthermore, results were considered robust if the optimal k solution re-appeared over different clustering algorithms and UMAP neighbor parameters. Finally, results were considered robust if the *SigClust* output rejected the null hypothesis of a single multivariate Gaussian for a vast majority of all repeat analysis runs. For different subject groupings, we also re-ran *reval* over autism-only and TD-only groupings separately. This allowed us to compare the robustness of clustering within autism or TD respectively. For reporting generalizability accuracy in the main results, we report the average accuracy over all 20 repeat analysis runs rather than reporting only one accuracy value from one of the seed states. Final subtype labels used in further downstream analyses are based on a majority vote across all 20 repeat runs over different random seeds.

For further tests of out-of-sample generalizability and replication of the clustering results, we used the CMI-HBN dataset under EO comprising H and γ and constructed a kNN classifier that learns the optimal k=2 k-means subtype solution. We then fit the CMI-HBN classifier to the IIT LAND replication dataset in order to predict subtype labels. The CMI-HBN classifier’s predicted subtype labels were then compared to labels from a k-means clustering of IIT LAND with k=2. A final generalizability accuracy can then be computed as the percentage overlap between the CMI-HBN classifier’s predicted labels and the actual IIT LAND clustering labels.

Once autism EEG-derived subtypes were identified, we tested for between-group differences between the autism subtypes and a typically-developing (TD) control group. For these analyses we utilized linear mixed effect models, computed per each electrode with the *lmer* function in the *lme4* R library. The dependent variable in these models were H or γ values. Subtype, age and the subtype*age interactions were modeled as fixed effects. Acquisition site was modeled as a random effect with random intercepts. Multiple comparisons correction was implemented with false discovery rate (FDR; q < 0.05). All between-group differences are descriptively shown in figures as standardized effect sizes, operationalized as *Cohen’s d* in standard deviation units.

### CMI-HBN behavioral data analyses

To conduct a comprehensive analysis of behavioral phenotypic data from the CMI-HBN dataset, we downloaded scores from all phenotypic tests and questionnaires in the CMI-HBN database (https://data.healthybrainnetwork.org/main.php). Downloaded data was then merged and filtered to only include autistic and TD individuals included in the EEG data analyses (n = 409). This dataset comprised a total of 14,195 variables. Next, some data-wrangling steps were taken to reduce the dimensionality of the dataset into a more manageable dataset with relevant test scores. First, columns specifying diagnosis codes, tracking and administrative information were dropped. Next, Social Responsiveness Scale (SRS) scores from the two forms (SRS_Pre (preschool-age) and SRS (school-age)) were combined into a single column. After these filtering and merging steps, the dataset was reduced to 9,620 variables. We then thresholded the dataset to only variables with data present >80% of sample (n=1,314 variables). Item-level variables were then discarded in favor of summary measures (e.g., totals, T-scores, standard scores, composites, and domain scores). Finally, we removed one datapoint that upon manual inspection revealed itself to be a very clear and large outlier. The cumulative result of all these data cleaning steps resulted in a final dataset with n=138 primary variables.

Analysis of behavioral differences was conducted for n=123 TD individuals and autistic individuals with consistent subtype labels over both EO and EC conditions (A1 n=108; A2 n=121). See Supplementary Table 12 for descriptive statistics of the final sample utilized in these analyses. For analysis of behavioral differences between groups we ran 3 linear models to specifically test for the full range of all pairwise group differences (i.e. TD vs A1, TD vs A2, A1 vs A2). These linear models were run via a permutation test whereby group labels were randomly permuted 10,000 times. This allowed for a non-parametric approach to estimate null distributions of the test statistic and to compute p-values that would be appropriate across all variables regardless of distributional assumptions. Multiple comparison control was achieved with FDR q<0.05. For some of the variables, when age significantly covaried with the dependent variable we used age as a covariate to control for in the linear model. Behavioral differences are described with standardized effect size (*Cohen’s d*) for each pairwise group comparison. We characterized 3 types of patterns of behavioral differences. First, case-control differences were identified as variables who had a conjunction of significant differences in the same directionality for both TD vs A1 and TD vs A2 comparisons. Otherwise, we identify ‘subtype-specific’ effects as variables that possessed a specific TD vs A1 or TD vs A2 difference or ‘subtype-differential’ effects as variables that showed a significant difference across all 3 pairwise group comparisons.

### CMI-HBN brain-behavior relationship analyses

For analysis of brain-behavioral relationships we used a multivariate association approach called partial least squares (PLS) correlation analysis^46^. Our PLS analyses are implemented with the *pls_analysis.m* function within the PLS MATLAB toolbox (https://www.rotman-baycrest.on.ca/index.php?section=84), using the Behavioral PLS method. PLS utilizes two matrices for testing multivariate brain-behavioral relationships - a brain matrix (i.e. rows correspond to subjects, columns correspond to H or γ features across all channels) and a behavioral matrix (i.e. rows correspond to subjects, columns correspond to specific behavioral variables). These matrices are stacked over groups and allow for behavioral saliences (indicating how important each behavioral variable is) to be output per each group. The main computational operation in PLS analysis is singular value decomposition (SVD) applied to the correlation matrix between brain and behavior matrices. This SVD is done to decompose multivariate brain-behavioral associations as a set of orthogonal latent variable pairs that explain covariance between brain and behavior matrices. There are 3 important outputs of PLS SVD on the brain-behavioral correlation - 1) the singular value δ, 2) a matrix V, interpreted as ‘brain saliences’, and 3) a matrix U, interpreted as ‘behavioral saliences’. The singular value δ can be used for hypothesis testing to identify statistically significant brain-behavioral relationships. To implement such hypothesis testing on each LV pair we use a permutation testing framework (10,000 permutations) whereby the singular value (δ) can be repeatedly recomputed when the rows of the matrices are randomly permuted to build a null distribution of singular values. Statistical significance is evaluated by counting the percentage of times amongst the 10,001 permutations that a singular value was greater than or equal to the observed singular value obtained in the unpermuted data. FDR control at q<0.05 is used to identify LV pairs that show a significant brain-behavioral relationship. In addition to the singular value (δ) from the SVD, we can also describe the percentage of covariance explained by each LV and this descriptive statistic is also reported for any significant LV pairs. Finally, brain and behavioral salience values are the other outputs of the SVD operation and allow for examination of which EEG channels (i.e. brain saliences) or behavioral variables (behavioral saliences) are most important for contributing to the brain-behavior association. To isolate specific EEG channels or ‘non-zero’ behavioral variables of most importance bootstrapping (10,000 bootstrap resamples) is used in the PLS analysis pipeline to compute brain bootstrap ratios (BSR) for EEG channels as well as 95% bootstrap confidence intervals for the behavioral variables. Brain BSRs can be thought of as pseudo-Z statistics and channels with BSR>2 are considered as important for driving the LV relationship. Behavioral variables with bootstrap confidence intervals that did not encompass a null correlation value of 0 were considered ‘non-zero’ variables that contribute most to the LV relationship.

PLS analyses were run separately on H and γ features and for EO and EC separately. These PLS analyses were run on all TD and autistic individuals that had consistent subtype labels over EO and EC conditions. Due to the fact that PLS cannot handle missing data, we implemented a procedure before running the analysis to check for missing data. Subjects with <80% of variables missing were removed from analysis. Additionally, we removed subjects above 15 years and below 6 years of age, due to the fact that many of these subjects had numerous missing variables. Thus, the final sample sizes for the PLS analyses were n=94 TD and n=176 autistic individuals (A1 n=87; A2 n=89). Of these remaining subjects the small amount of missing data was handled with kNN imputation (k=10). All n=138 variables from the behavioral analyses were used in the PLS analysis. However, we added age as a final variable for the PLS analyses, raising the total variables in the PLS analysis to n=139. See Supplementary Table 12 for descriptive statistics of the final sample utilized in these analyses.

In addition to isolating PLS LVs that were statistically significant, we also evaluated the behavioral salience vector outputs of the PLS analyses between groups and also between EO and EC conditions. To evaluate within-group robustness of brain-behavioral relationships we computed the correlation between behavioral salience vectors of the same group between EO and EC analyses. Significant correlations indicated a high-level of within-group robustness of brain-behavioral associations, signaling the relationships are invariant to changes in EO versus EC conditions. In addition to testing within-group robustness, we also tested for between-group dissimilarity in brain-behavioral association via computing the correlation between behavioral salience vectors for all pairwise group comparisons within a particular EO or EC analysis. Significant correlations on these tests indicate that brain-behavioral relationships are significantly similar between-groups. In contrast, non-significant between-group correlations retain the null hypothesis that brain-behavioral relationships are not significantly similar between-groups. Finally, to visualize the most important behavioral variables and domain categories with non-zero relationships we generated word clouds of the 13 behavioral domains where the size of the word was modulated by the weighted average behavioral salience correlation of the non-zero variables. The weight used in these weighted averages was defined as the percentage of variables within a specific domain category that had a non-zero relationship.

### In-silico modeling of LFP and EEG data

The *in-silico* modeling simulates a recurrently connected network model of leaky-integrate-and-fire (LIF) pyramidal (excitatory) and interneuron (inhibitory) spiking neurons^13^ and computes highly realistic approximations (proxies) of the extracellular potentials (LFP, EEG) that these neurons would generate^19,47^. This recurrent spiking network model represents a relatively standard model of a recurrent cortical circuit of recurrently connected E and I populations that receive external inputs. The network structure and parameter range are similar to those used in past work^11,19,48^ (Supplementary Table 13-14). The network is composed of 5000 neurons, of which 4000 are excitatory (i.e. they form AMPA-like excitatory synapses with other neurons) and 1000 inhibitory (forming GABA-like synapses). Neurons are randomly connected with a connection probability between each pair of neurons of 0.2. Both populations receive as external depolarizing inputs two sets of spike trains. The first, and larger, input is a set of spikes generated with a time-independent Poisson process whose mean value *ν* is varied across the simulations performed here and was used to regulate the average level of depolarization. The larger *ν* is, the more the neurons are depolarized on average at any instant of time. The second set of input spike trains are generated with a time-dependent firing rate obeying a rectified Ornstein–Uhlenbeck (OU) process, allowing the network to experience low-frequency input variability such as that which could reflect large-scale brain state changes. This second time dependent process had parameters that were kept constant across all simulations, that generated slow fluctuations with most power below 10 Hz, and that made it much smaller in mean amplitude (7.5-20 times) than the time-independent one.

In a first set of simulations, reported in Fig. 1, we changed across simulations two microscopic parameters: The first is a ratio called *g* operationalized as the ratio between excitatory AMPA (*g_EE_*) and inhibitory GABA (*g_IE_*) synaptic conductances onto E neurons (i.e. *g* =*g_EE_*/*g_IE_*). Following Trakoshis et al.,^11^ the *g* ratio was modulated by keeping *g_EE_* fixed and changing *g_IE_*. We also rescaled *g_II_* proportionally to the change of *g_IE_* to rescale inhibition strength uniformly across all neuron types. The *g* ratio was varied across simulations between 4-14 times greater inhibition relative to excitation. The second parameter that was systematically varied is the external input rate to the network, called ν, expressed in units of spikes per second per neuron indicating the average number of spikes per unit time sent by each input neuron. The external input rate, ν, was modulated between 1.5 to 4 input spikes per second per neuron. Both of these parameters spanned ranges that reproduce well both spontaneous and stimulus-evoked extracellular potentials and firing regimes in the cortex^11,14,20^. The network had irregular (Poisson-like) firing for all parameters considered, and when increasing *ν* or *g* it transitions from asynchronous irregular (AI, with lower correlations between neurons) to synchronous irregular (SI, with higher correlations between neurons) firing regimes (Fig. 1b-c). To assess the network’s dynamical regimes, we computed three metrics: the mean firing rate (FR) of excitatory neurons, the coefficient of variation (CV) of inter-spike intervals (ISI) from excitatory neuron spikes, and the mean pairwise correlation coefficient among excitatory neurons. Based on the criteria defined by Kumar and colleagues^49^, we classified the parameter space into asynchronous irregular (AI) and synchronous irregular (SI) regimes. We defined the mean firing rate of E and I neurons in the model or their weighted average *FR_AVG_* = 0.8\**FR_E_* + 0.2\**FR_I_* as a measure of excitability of the network.

In a second set of simulations, we mimicked the chemogenetic manipulations of hSYN-hM4Di, CamkII-hM3Dq, and PV-hM4Di experiments. For the hSYN-hM4Di experiment, we simultaneously varied the resting membrane potential (*E_l_*) of both E and I neurons between −70 mV (our reference value) and −85 mV, thus inducing hyperpolarization when *E_l_* becomes more negative. We also reduced synaptic strength across all connections to emulate synaptic silencing of the hSYN-hM4Di manipulation^26^. Synaptic strength was reduced proportionally to the decrease of *E_l_* (by 0.05 nS for a decrease of 1 mV in *E_l_*). To simulate the effects of the CamkII-hM3Dq experiment we varied the resting membrane potential (*E_l_*) of E neurons between −70 mV (our reference value) and −66 mV, thus inducing depolarization when making *E_l_* less negative. To simulate the effects of the PV-hM4Di experiment we varied the resting potential (*E_l_*) of I neurons between −70 mV (our reference value) and −71.5 mV. We also reduced synaptic strength across all I (g_IE_ and g_II_) connections to emulate synaptic silencing of the PV-hM4Di manipulation^26^. Synaptic strength was reduced proportionally to the decrease of *E_l_* (by 0.3 nS for a decrease of 1 mV in *E_l_*).

In a final set of control simulations, we changed the number of inhibitory neurons while keeping the excitatory population fixed (Supplementary Fig. 3). Because altering the size of the inhibitory population changes the total amount of inhibitory input a neuron would receive, we rescaled the inhibitory synaptic conductance proportionally to the change in inhibitory population size. This ensured that each neuron experienced, on average, the same net inhibitory drive across all tested network configurations. All other network parameters and simulation details were identical to those of the default model described above for the first set of simulations. We found that in this case the firing rate of E and I neurons varied similarly with the parameters as the original network because what determines the average firing rate of a neuron is its average synaptic drive, which relates to the product of the average synaptic conductance and the number of presynaptic neurons multiplied by their firing rate. This implies that similar dynamics can be reached varying either the number of I neurons or the average synaptic strength (see Supplementary Fig. 3).

Across all simulations, the LFP is computed as the sum of absolute values of AMPA and GABA postsynaptic currents on excitatory cells^14,47^. This simple estimation of LFPs was shown to capture more than 90% of variance of both experimental data recorded from cortical field potentials and of simulated LFPs that would have been produced by the same spiking activity within a network of 3D neurons with highly realistic morphology^14,20,47^. Using simulated AMPA and GABA currents from the model, we also computed the ERWS2 EEG proxy as described in prior work^19^. The ERWS2 EEG proxy used here accounts for approximately 95% of the variance of ground truth EEG signal produced by the same spiking activity within a network of 3D neurons with highly realistic morphology^19^. Using both simulated LFP and EEG proxy data from the model, we computed H identically to how H was computed in human EEG data and the 1/f slope as computed by Gao and colleagues^10^ using the FOOOF algorithm^6^ with the following parameters: max_n_peaks = 2, peak_width_limits = [2, 8], peak_threshold = 1, and mode = “fixed”. γ-band power was calculated as the integral of the power spectral density within the 30–50 Hz frequency range to match the frequency range used in human CMI-HBN EEG data.

To assess how variation in the parameters relates to LFP/EEG features, we computed Pearson correlations between each parameter and the LFP measures (H, 1/f slope, total power, and γ-band power). We then evaluated how well these measures could predict firing rate by training linear models (*fitlm.m* function in MATLAB) that used either individual measures or pairs of measures (bivariate regression) as predictors of firing rate. Model performance was quantified as the mean and standard deviation of the Pearson correlation between predicted and actual firing rates across 5 cross-validation folds.

### In-vivo chemogenetic experiments altering excitation-inhibition balance

All in-vivo studies in mice were conducted in accordance with the Italian law (DL 26/214, EU 63/2010 Ministero della Sanità, Roma) and the recommendations in the Guide for the Care and Use of Laboratory Animals of the National Institutes of Health. Animal research protocols were also reviewed and consented to by the animal care committee of the Istituto Italiano di Tecnologia and University of Trento. Within these approved research protocols, we estimated sample sizes based on previously published effect sizes in the literature or via internal pilot studies.

Experimental procedures for viral expression and chemogenetic manipulations have been described in greater detail elsewhere^25^. Briefly, adult male C57Bl6/J (hSYN-hM4Di and CamkII-hM3D(Gq) manipulations) or Pvalb^tm1(cre)Arbr^ (PV::Cre; Jackson Laboratories; Bar Harbor, ME, USA) mice (mean age = 17 weeks, SD age = 6 weeks) old were anesthetized with isoflurane (isoflurane 4%) and head-fixed in a mouse stereotaxic apparatus (isoflurane 2%, Stoelting). Viral injections were performed with a Hamilton syringe mounted on Nanoliter Syringe Pump with controller (KD Scientific), at a speed of 0.05 μl/min, followed by a 5–10 min waiting period, to avoid backflow of viral solution and unspecific labeling. Viral suspensions were injected bilaterally in the mouse medial PFC using the following coordinates, expressed in millimeters from bregma: 1.7 AP, −0.3 ML, −1.7 DV. To account for the different transfection and tropism capabilities across viral vector serotypes^50^, different injection volumes determined from pilot studies were used for each experiment to achieve a comparable anatomical spread of the employed DREADD receptor within the mouse PFC (Supplementary Fig. 4). The targeted regions included all major PFC subdivisions: prelimbic (PL), infralimbic (IL), and anterior cingulate cortex (ACC) as per the broad medial prefrontal cortex definition by Vogt and Paxinos^51^.

In all our *in-vivo* experiments we used experimental (DREADD) and SHAM control groups. For the DREADD pan-neuronal silencing experiment (hSYN-hM4Di), hM4Di DREADD was transduced using an AAV8-hSyn-hM4D(Gi)-mCherry construct. Age and genotyped matched control animals (SHAM) were injected with a AAV8-hSyn-GFP virus (www.addgene.com). These viral suspensions were injected using a 1µL bilateral injection volume in n = 5 hM4Di DREADD and n = 5 SHAM mice, respectively. For the CamkII-hM3Dq DREADD excitation experiment, CamkII-hM3D(Gq) DREADD was transduced using an AAV9-CamkII-hM3D(Gq)-mCherry construct which we injected bilaterally into the PFC as described above at a volume of 250 nL per hemisphere. Age and genotyped matched control animals (SHAM) for this experiment underwent a surgical procedure during which injection needles were inserted into parenchymal cerebral tissue at the correct stereotaxic coordinates, but no viral suspension was administered. This experimental cohort was composed of n = 5 CamkII-hM3D(Gq) DREADD mice, and n = 8 SHAM. Similarly, for the PV-hM4Di experiment, 400 nl of AAV8-hSyn-DIO-hM4Di-mCherry viral suspension were injected bilaterally in the same coordinates of n=3 mice. As in the CamkII-hM3D(Gq) manipulation, control animals (SHAM) (n=4) underwent surgeries without viral infusion. Prior to each DREADD experiment, we waited at least 3 weeks to allow for maximal viral expression. For all experiments DREADD activation was carried out upon injection of CNO in both SHAM and DREADD groups. In all our electrophysiological recordings we acquired a pre-CNO baseline followed by a post-CNO active window. CNO was only administered on test day to avoid plasticity related effects due to repeated administrations.

All *in-vivo* experiment electrophysiological recordings were carried out using established light sedation protocols. The use of light sedation was primarily aimed at obtaining a stable brain state throughout the duration of the manipulation, comparable to quiet wakefulness in human recordings. This approach replicates conditions commonly employed in mouse resting-state fMRI studies, for which these manipulations were originally developed and validated^25^. A key advantage of light sedation as opposed to measurements under head-fixed awake conditions is the ability to minimize fluctuations in arousal and spontaneous behaviors, thereby ensuring a consistent and interpretable brain state for measuring resting neural activity. This is particularly important when assessing subtle and prolonged neuromodulatory effects such as those induced by chemogenetic manipulations. The hM4Di silencing experiment was conducted under halothane sedation^25,52^. However, due to the commercial discontinuation of halothane, subsequent CamkII-hM3D(Gq) and PV-hM4Di experiments were carried out using a medetomidine-isoflurane combination (“med-iso”). The med-iso combination is now considered the gold standard for mapping of resting brain activity in rodents^53,54^. Importantly, all experimental and control groups were sedated using the same protocol within each experiment, ensuring that statistical comparisons were not confounded by differences in anesthetic regimen. All inferences were based on within-experiment DREADD vs SHAM comparisons under identical sedation regimes.

For hSYN-hM4Di experiments, electrophysiological recordings were carried out using animal preparation and sedation regimes employed in prior studies^25,52,55^. Surgery and preparations were made while mice were anesthetized with isoflurane (4% induction), intubated, artificially ventilated (2% maintenance), and head-fixed in a stereotaxic apparatus (Stoelting). The tail vein was cannulated for clozapine-N-oxide (CNO) injection (2 mg/kg, administered to both control and experimental cohort). To ensure maximal consistency between viral injections and recording site, the skull surface was exposed and an insertion hole was gently drilled through the skull corresponding to the location of prior viral injection point. A 16-channel linear probe (Neuronexus, USA) was next inserted through the overlying dura mater by a microdrive array system (Kopf Instruments, Germany) at an insertion rate of 1 μm/min to reach the same stereotaxic coordinates employed for viral injection. Electrode insertion was performed in 3 steps of similar length, with a 20-minute waiting period in between to allow the tissue to reposition after each insertion step. During the last step of electrode positioning, isoflurane was discontinued and replaced by halothane at a maintenance level of 0.75%. Electrophysiological data acquisition commenced 1 hour after isoflurane cessation. Such transition time was required to ensure complete washout of isoflurane anesthesia and avoid residual burst-suppressing activity associated with extended exposure to deep anesthetic levels.

CamkII-hM3D(Gq) and PV-hM4Di electrophysiological acquisitions were carried out using a combination of low dose isoflurane and medetomidine^53^. Briefly, surgery and preparations were made while animals were anesthetized with isoflurane (4% induction, 2% maintenance), intubated and head fixed on the stereotaxic frame. The tail vein was cannulated for medetomidine infusion. After craniotomy, electrode coordinates and insertion were performed as described before. The tail vein was cannulated for posterior medetomidine infusion, and a second cannula was placed intraperitoneally for CNO administration (0.5 mg/kg for CamkII-hM3D(Gq), and 2 mg/kg for PV-hM4Di experiments, with matched dosage for the corresponding controls). Temperature was constantly monitored and maintained to 36.5+-0.5 °C. Craniotomy procedures, and electrode insertion were performed as described before. Following the last step of the electrode insertion, sedation via a combination of medetomidine and isoflurane commenced. An initial intravenous bolus of 0.05 mg/kg of medetomidine was injected, and isoflurane was lowered to 1%. After 5 minutes, isoflurane level was lowered further to 0.3-0.5% and a constant infusion of medetomidine (0.1 mg/kg/h) was applied for maintenance until the end of the acquisition. For all electrophysiological experiments, neural activity was recorded after electrode positioning in consecutive 5-minute time bins to cover a 15 min pre-injection time window, and a 60 min post CNO timeframe. Signals were amplified using an RHD 2000 amplifier system (Intan Technologies, RHD Recording Controller Software, v2.09) to acquire electrophysiological data at a sampling rate of 20 kHz.

To compute the LFP signal, raw extracellular recordings were first downsampled to 4 kHz, then band-pass filtered to 0.1-250 Hz using a two-step procedure^56^. Briefly, raw time-series were first low-pass filtered using a 4th order Butterworth filter with a cut-off frequency of 1 kHz. The resulting timeseries were next downsampled to 2 kHz, then again filtered using a Kaiser window filter between 0.1 Hz to 250 Hz (with a sharp transition bandwidth of 1 Hz, passband ripple of 0.01 dB and a stop band attenuation of 60 dB) and then resampled at 1 kHz. Filtering was applied both forward and backward to remove filtering phase transitions lags.

Multi-unit activity (MUA) was computed following the procedure described by Belitski and colleagues^56^. A high-pass filter was applied to the extracellular signal (4th order Butterworth filter with cut off frequency over 100Hz), followed by a band-pass filter between 400 and 3000 Hz using a Kaiser window filter (with transition band of 50 Hz, stopband attenuation of 60 dB, and passband ripple of 0.01 dB). Events over a threshold corresponding to 4-times the median of the signal of all traces (baseline and post injection combined), divided by 0.6745 ^57^ were counted as neuronal action potentials (spikes). For the final count, spikes were considered to be biologically plausible, and therefore retained, only if occurring more than 1 ms apart.

### DREADD LFP data analysis

Preprocessed LFP and MUA data from the DREADD experiments were cut into 4-second segments, with approximately 1 minute of separation between each segment. Given the high level of correlation between the 16 channels of recorded LFP data, we used principal component analysis as a dimensionality reduction technique to capture the vast majority of shared variance (e.g., 95-99%) across channels into one variable (PC1). PC1 was then utilized as input for computing H in a manner identical to how H was computed for human EEG data. The DREADD experiments consisted of 3 phases, called baseline, transition, and treatment. The baseline phase was a 15-minute time period before the drug injection. The transition phase was defined here as a subsequent 20-minute period post-injection when the drug begins to exert its effect before reaching full pharmacokinetic equilibrations^25,58^. The treatment phase followed the transition phase (duration 25 minutes) and is considered as the time period where the drug is predicted to exert its maximal effect. Before hypothesis test statistical modeling, the mean and standard deviation (SD) of H for each mouse was computed within the baseline period and then used to normalize all H values, computed as in human EEG, across the entire experiment as a baseline normalization. This allows the dependent variable in our statistical model to be baseline normalized H, quantifying H in terms of change from baseline average H in units of SD. The same baseline normalization was carried out for the analysis of the 1/f slope, computed between 20-90 Hz using the FOOOF algorithm^6^ with the following parameters (max_n_peaks: 2; peak_width_limits: [2,8]; peak_threshold: 1; mode: “fixed”), and for the integrated spectral power, defined as the sum of the LFP power spectrum across all frequencies (0-90 Hz), and γ power computed as the sum of LFP power spectrum across 30-50 Hz, as with EEG human dataset. We verified that the results of the paper were robust to variations of the FOOOF free parameters in a broad range.

Statistical modeling for hypothesis testing of changes over time on LFP H data, 1/f slope, and integrated spectral power for the DREADD experiments was implemented using linear mixed effect models, implemented using *fitlme.m* function of MATLAB. The dependent variable in the model was baseline normalized. Fixed effects were group (DREADD) and condition (baseline, transition, treatment) and their interaction. Random effects in the model were modeled with random intercepts for each mouse. The same hypothesis testing approach with linear mixed effect models was used for MUA spiking activity data. Regression analyses were conducted using the MATLAB *fitrobust.m* function with the Huber loss to mitigate the influence of outliers. A 10-fold cross-validation procedure was applied to the dataset, and Pearson correlation coefficients were computed to assess the predictive performance between the actual and predicted firing rates.

To compare the predictive performance of the different models (H, 1/f slope, γ, H+γ, total power and simulation-based predictors), we adopted a bootstrap-based evaluation framework. Each model was trained and evaluated on 1,000 bootstrapped datasets constructed by resampling pooled single-trial data from the treatment phase of the hSyn-hM4Di, CaMKIIα-hM3D(Gq), and PV-hM4Di experiments. For each bootstrap iteration, model performance was quantified as the correlation between predicted and observed baseline-standardized FR, yielding an empirical distribution of performance values for each predictor. From these distributions, we estimated the mean and variance of model performance. Statistical comparisons between models were then performed using pairwise z-tests on the difference between mean performance values, with the z-statistic computed using the pooled standard error derived from the bootstrap distributions. Corresponding two-sided p-values were obtained from the standard normal distribution. To estimate firing rates using simulation-derived weights, we utilized the mean weights obtained from the 10-fold cross-validated models in conjunction with the corresponding 10-fold cross-validation data partitions.

## Data availability

CMI-HBN data can be found at http://fcon_1000.projects.nitrc.org/indi/cmi_healthy_brain_network/. IIT LAND data can be found at https://gitlab.iit.it/bmp006-public/LAND_IIT/cmi_eeg_h-gamma. Electrophysiological data of the DREADD manipulations in the mouse PFC can be found in the following repositories: hSYN-hM4Di (10.5281/zenodo.17991893), CamkII-hM3Dq (10.5281/zenodo.18010569 and 10.5281/zenodo.18011691) and PV-hM4Di (10.5281/zenodo.18004527).

## Code availability

The *reval* Python library can be found on GitHub (https://github.com/IIT-LAND/reval_clustering) and the documentation can be found at https://reval.readthedocs.io. Analysis code for the study is available on our GitHub repo (https://gitlab.iit.it/bmp006-public/LAND_IIT/cmi_eeg_h-gamma). The code for the simulation of point-neuron networks is available in our GitHub repository (https://github.com/panzerilab/Electrophysiologically-defined-excitation-inhibition-autism-neurosubtypes).

## Acknowledgments

This project was supported by funding from the Simons Foundation for Autism Research Initiative (SFARI; grant number 982347) to MVL, AG, and SP and from the European Research Council (ERC) under the European Union’s Horizon 2020 or Horizon Europe research and innovation programmes (grant agreement numbers 755816 (AUTISMS) and 101087263, (AUTISMS-3D) to MVL; grant agreement numbers 802371 (DISCONN) and 101125054 (BRAINAMICS) to AG).

## Author Contributions

Conceptualization: MVL, NB, AG, SP. Methodology: MVL, NB, AV, VM, GML, DSY, AG, SP. Formal analysis: MVL, NB, AV, VM, GML, GM, SBM, SP. Investigation: MVL, NB, EMB, SD, MF, OJL, IS, MR, AB, PV, NP, KP, CB, SC, EC, DSY, GML, GM, SBM. Writing - original draft preparation: MVL, NB, GM, DSY, GML, SBM, DB, AV, VM, PMC, AG, SP. Writing - review and editing: MVL, NB, GML, GM, SBM, DSY, AV, DB, VM, PMC, AG, SP. Visualization: MVL, NB. Supervision: MVL, AG, SP. Project administration: MVL, AG, SP. Funding acquisition: MVL, AG, SP.

---

**Extended Data Fig. 1:**
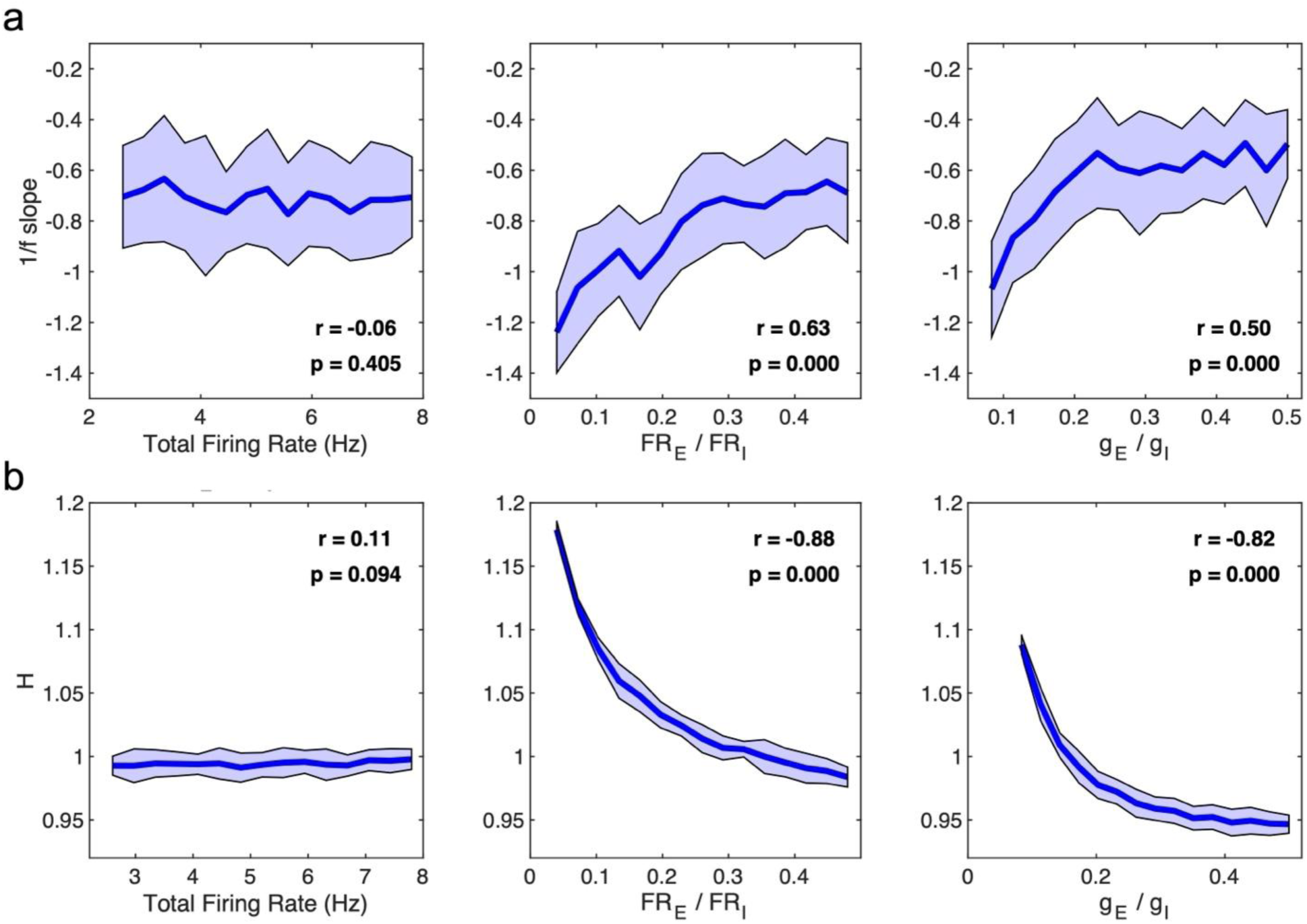
Analysis of E:I LFP proxies from the uncoupled model from Gao et al.,^10^. This figure presents the results of the analysis of how the LFP proxies of E:I imbalance depend on neural parameters in the uncoupled excitation-inhibition model described by Gao et al.,^10^. In this model, E and I spikes are generated at random times (homogeneous Poisson process) with a given mean firing rate of each cell type (FR_E_ for E neurons; FR_I_ for I neurons). Post-synaptic responses are created exactly as in Gao et al.,^10^ by convolving the spike trains with a beta function. The beta function has parameters τ rise= 0.1ms and 0.5 ms, and τ decay = 2ms and 10 ms for AMPA and GABA synapses respectively. The beta function is multiplied by a synaptic efficacy parameter g_E_ for E neurons and g_I_ for I neurons, and the LFP is computed as a sum of all synaptic currents, exactly as in Gao et al.,^10^. **a,** Plots showing 1/f slope on the y-axis, computed as linear interpolation as in Gao et al.,^10^ from the LFP in the 30-70 Hz range. The plots show the dependence of 1/f slope on either the grand average firing rate defined as the ratio of the firing of all E and I neurons (left), the ratio of E vs I firing rates (center), or the ratio of synaptic efficacies (g_E_/g_I_; right). **b,** Similar plots as panel **a**, but for H plotted on the y-axis rather than 1/f slope. In all panels, the thick line plots the mean and the shaded area shows the SD over n=15 simulations (10 seconds of simulated activity per simulation) for each parameter value. Each plot also shows in text the Pearson’s r and corresponding p-value for the association between plotted variables. Both H and 1/f slope in this model correlate tightly with variations in E:I conductance or firing rate ratio, but not with the total firing rate. In contrast, in both our in-silico coupled E:I network model and in-vivo mouse data, H and the 1/f slope correlate with the total firing rate.

**Extended Data Fig. 2:**
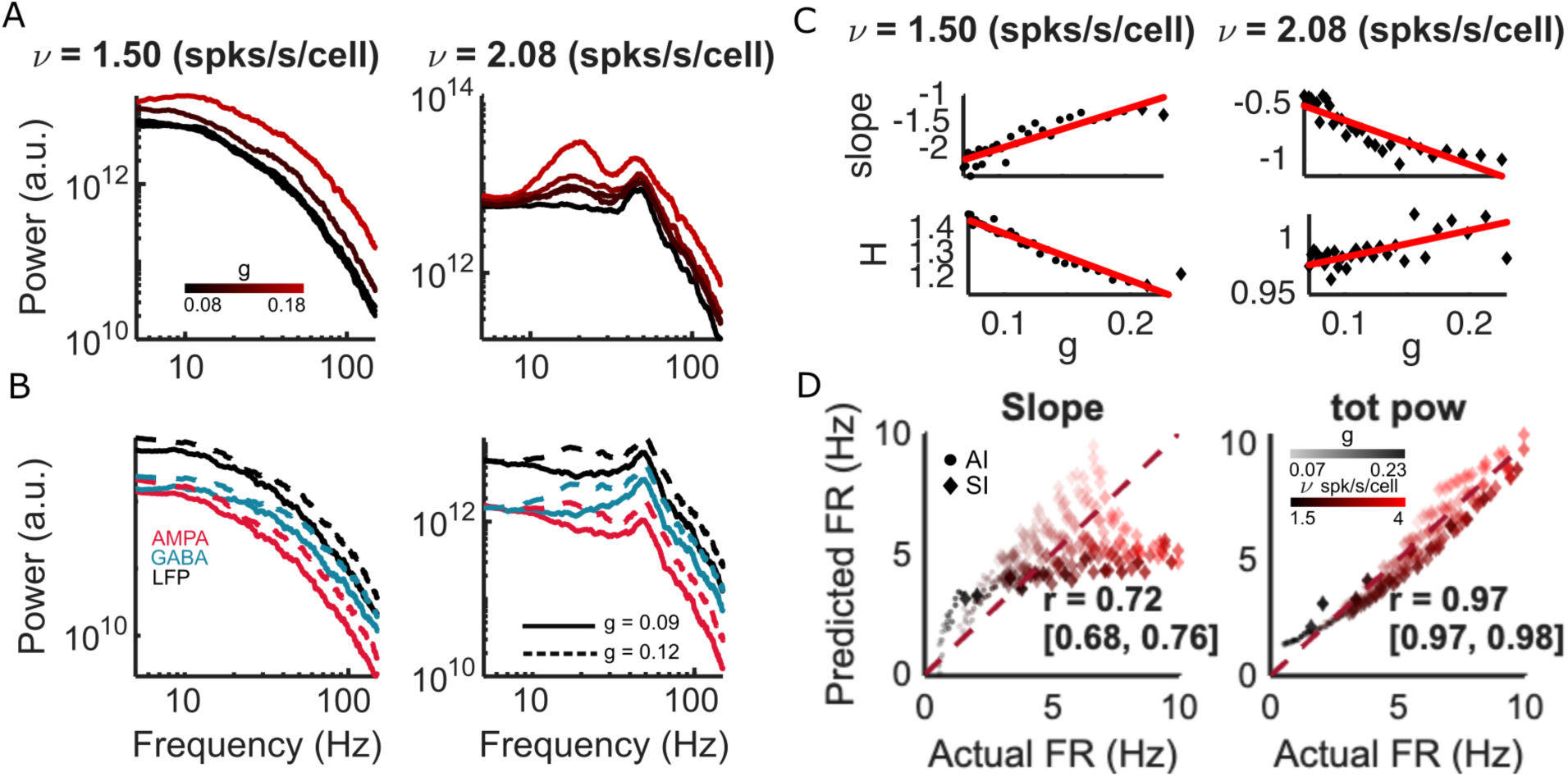
Further simulations of in-silico LFP spectral properties. **a**, Plot of spectral LFP power as function of frequency obtained from simulations with different values of g and ν parameters (indicated with text in panel title or color scale). The left panel plots an example in the asynchronous irregular (AI) regime (i.e. ν = 1.5 spikes per second per neuron) while the right panel plots an example in the synchronous irregular (SI) regime (i.e. ν = 2.08 spikes per second per neuron). **b,** Plot of spectral power of the AMPA (red) and GABA (blue) synaptic currents to E neurons and of their sum (to emulate LFP, black line) as function of frequency obtained from simulations with different values of g and ν parameters. This shows that both E and I currents spectra - and thus the LFP spectrum become flatter (i.e. proportionally more high frequency power) when increasing g or ν. **c,** Scatterplots showing how H and 1/f slope vary as a function of the g parameter with two different values of input ν. Each dot (AI) or diamond (SI) represents the outcome of an individual simulation with the considered parameter values. The least squares best fit lines are plotted in red. **d,** Plot across simulations of the true actual average firing rate (x-axis) against predicted firing rates (y-axis) by a linear model using only 1/f slope (left), or total power (right) as predictor variables. The Pearson’s r and 95% CIs computed from 5-fold cross-validation are shown in text. The diagonal line is also plotted to show what would be perfect prediction accuracy. Dots versus diamonds represent simulations within the SI and AI firing regimes respectively. 1/f slope does not correlate strongly with g (r = 0.187), but does correlate with v (r = 0.868). Similarly, total power is not correlated with g (r = 0.113), but is correlated with ν (r = 0.871) (see Supplementary Τable 1).

**Extended Data Fig. 3:**
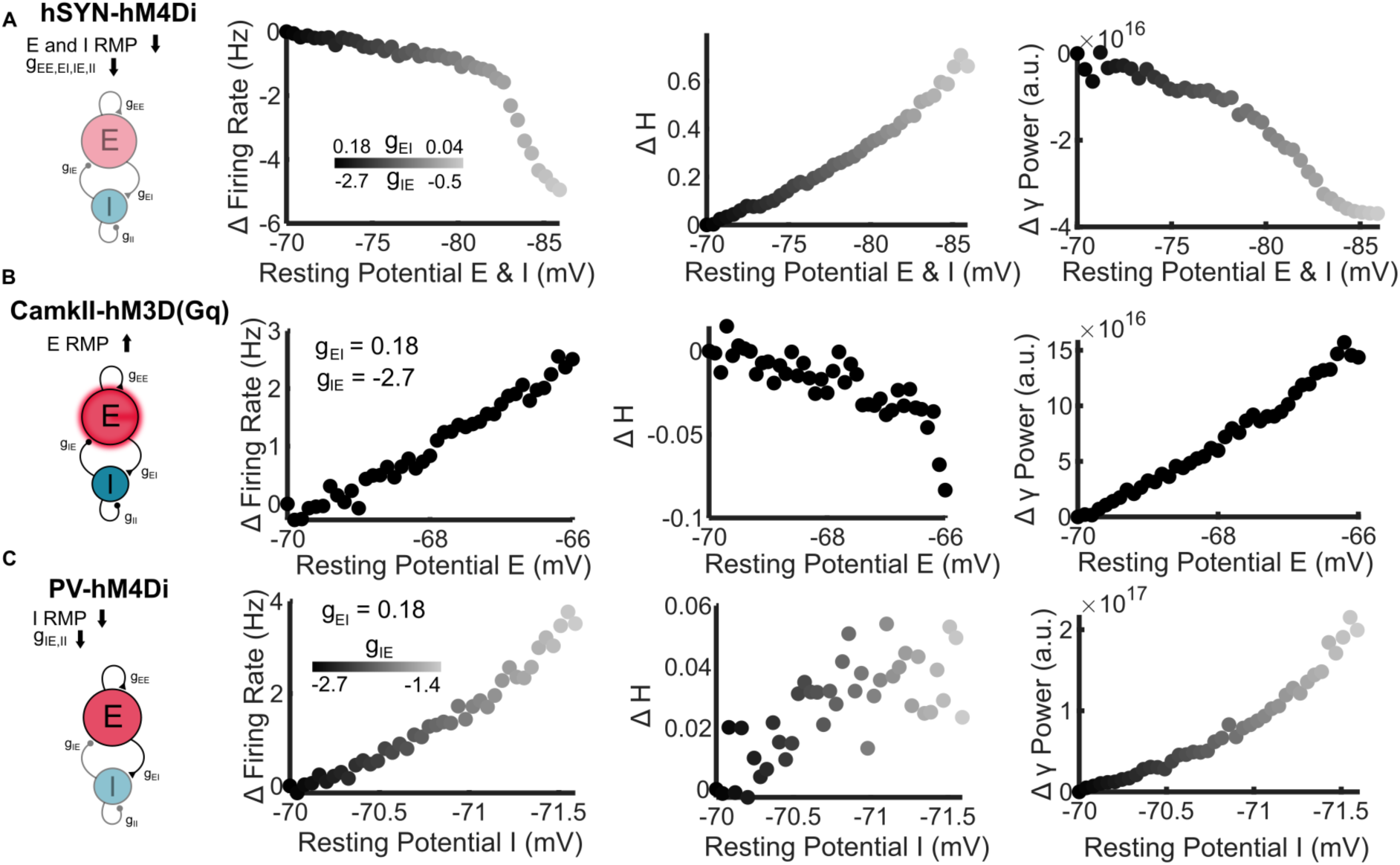
Simulated effect of chemogenetic manipulations with recurrent E-I networks. Here we simulated the effect of hSYN-hM4Di (silencing; panel **a**), CaMKII-hM3Dq (excitation of E neurons; panel **b**) and PV-hM4Di (inhibition of PV neurons; panel **c**) DREADD manipulations that we implemented in subsequent in-vivo validation experiments. On the far left are schematics of the performed manipulation, using grey levels and transparency to indicate which microscopic neural component (i.e. excitability of individual cells within a population, or synaptic strength of a population) was manipulated. Columns 2–4 show scatterplots from simulations in which either synaptic conductances (g) or resting membrane potential (E_l_) were varied to decrease (hSYN-hM4Di) or increase (CaMKII-hM3Dq and PV-hM4Di) neuronal excitability. Changes in excitability were primarily implemented as shifts in resting membrane potential, as indicated on the x-axis (see Methods). Plots show the change from baseline (Δ) in mean firing rate, the Hurst exponent (H), and γ-band power as a function of the manipulated parameter. Each dot represents the outcome of a single simulation at the corresponding parameter value. Abbreviations: RMP, resting membrane potential.

**Extended Data Fig. 4:**
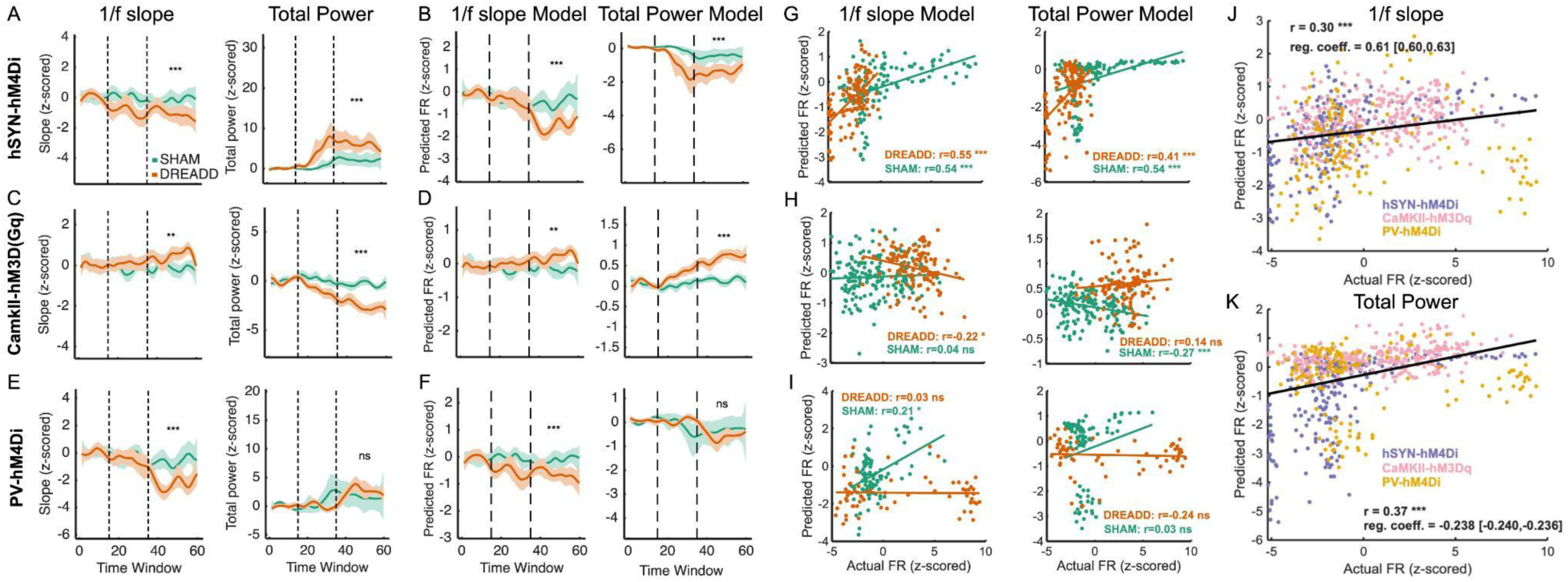
Further analyses of in-vivo chemogenetic manipulations that alter network excitability, using 1/f slope or total power as dependent variables. Panels a-f show baseline normalized trajectories of change computed over sliding time windows (x-axis) with each time window representing a 4 second segment of time and with each window separated by 1 minute. **a-b,** Results for the hSYN-hM4Di experiment. **c-d,** Results for the CamkII-hM3Dq experiment. **e-f,** Results for the PV-hM4Di experiment. Panels titled ‘1/f slope’ show the trajectories of 1/f slope on the y-axis. Panels titled ‘Total power’ have total broadband power on the y-axis. Panels titled ‘1/f slope Model’ or ‘Total Power Model’ show predicted firing rates using either 1/f slope (left) or total broadband power (right) as predictor variables. Panels **g-i,** show scatterplots of predicted firing rates (y-axis) against actual firing rates (x-axis) for all experiments (**g**, hSYN-hM4Di; **h**, CamkII-hM3Dq; **i**, PV-hM4Di). Each dot represents a prediction for each timepoint and animal. The solid line indicates the robust fit, with the corresponding Pearson correlation coefficient (r) and p-value reported. **j**, Scatterplot of actual firing rates (x-axis) against predicted firing rate (y-axis) using 1/f slope as a predictor, pooled over all trials and animals from all 3 experiments. **k**, Same as panel j, but using predicted firing rate (y-axis) with total broadband power as a predictor.

**Extended Data Fig. 5:**
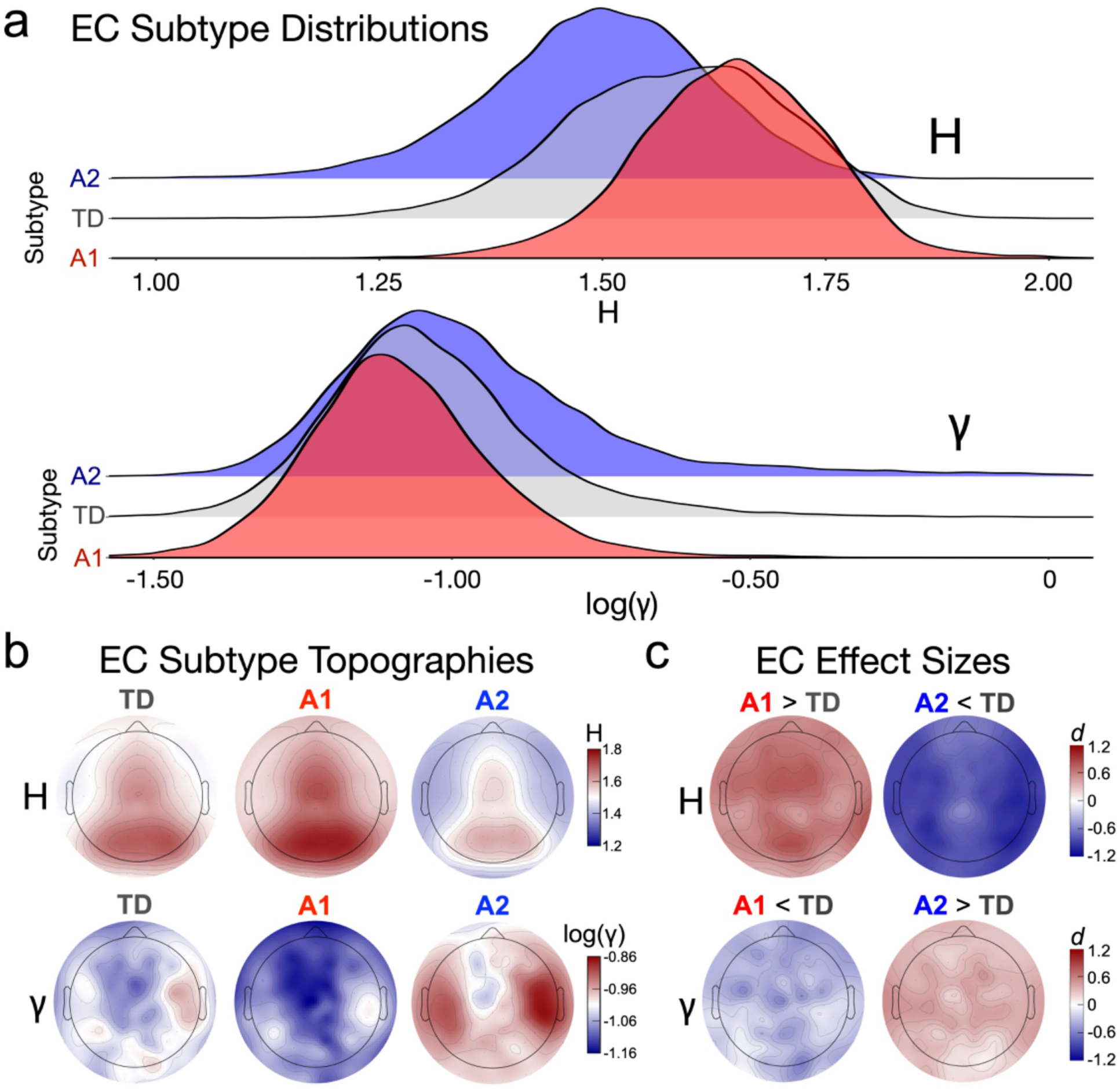
Subtype results for EC in the CMI-HBN dataset. **a**, Density plots of H (top) and γ (bottom) for the CMI-HBN dataset EC condition. Plots show the TD (gray), A1 (red), and A2 (blue) subtypes separately. **b,** Scalp topographies for H (top) and γ (bottom) during EC for TD, A1, and A2 subtypes. **c,** Standardized effect size (Cohen’s d) scalp topographies for EC H (top) and γ (bottom) for A1 vs TD (left) and A2 vs TD (right) comparisons in the CMI-HBN dataset.

**Extended Data Fig. 6:**
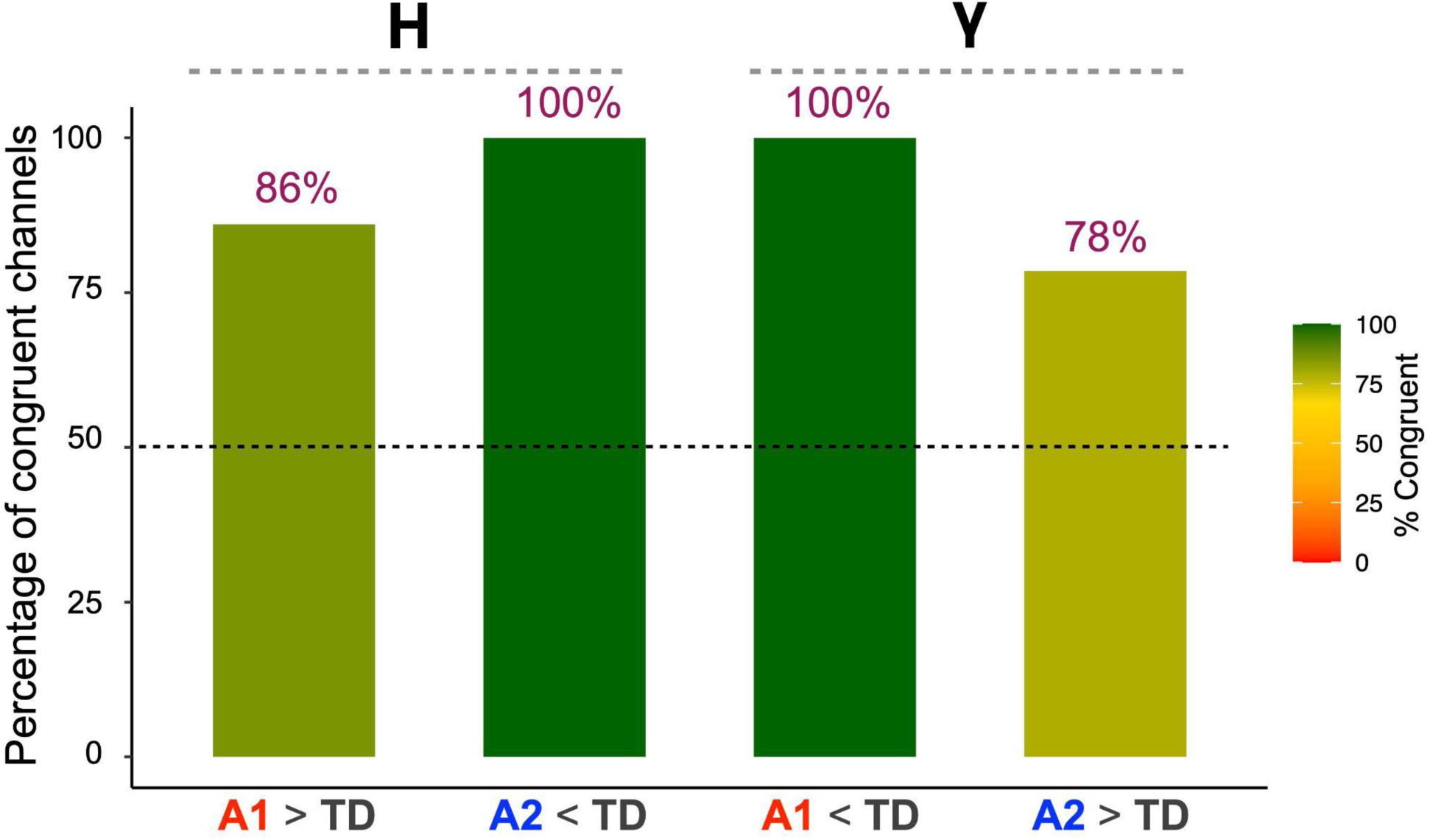
Effect size directionality replication in IIT LAND. Bar plot of the percentage of EEG channels where the directionality of the effect sizes were congruent between CMI-HBN EO and IIT LAND Inscapes.

**Extended Data Fig. 7:**
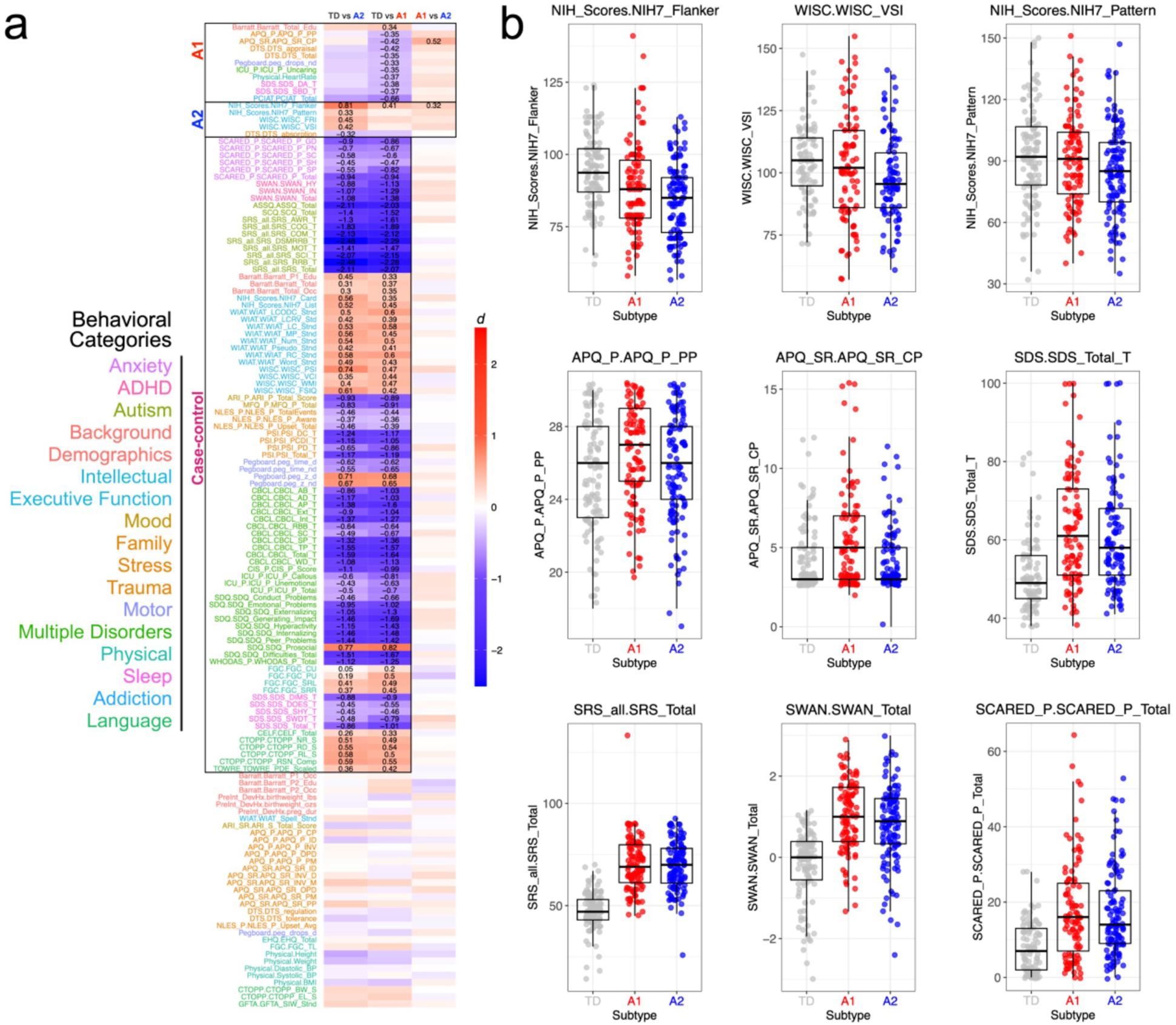
Phenotypic differences between groups in the CMI-HBN dataset. **a**, Heatmap of standardized effect sizes (Cohen’s d) of 138 variables across 13 domains (domain colors on the left). The 3 group comparisons are noted at the top of the heatmap. Specific effect sizes shown in numbers within each cell denote the comparisons that were significant after FDR q<0.05 correction for multiple comparisons. Variables are sorted from top to bottom and labeled as A1, A2, or case-control differences. A1 effects are variables where there was a significant difference between TD vs A1 and/or an A1 vs A2 difference, but where the TD vs A2 comparison was not significant. A2 effects are variables where there was a significant difference between TD vs A2 and/or A1 vs A2, but where the TD vs A1 comparison was not significant. Finally, case-control differences are variables whereby both TD vs A1 and TD vs A2 are significant, but A1 vs A2 is not significant. **b,** Examples of specific variables as scatter-boxplots.

## Supplementary Material

### Supplementary Results: Further analysis of the uncoupled model by Gao et al

Seminal work by Gao and colleagues^10^ used an uncoupled E:I model and proposed that the LFP 1/f slope (which captures scale-free properties similar to the fractal properties also captured by H) reflects the ratio *g* of E:I synaptic conductances. However, we show (Extended Data Fig. 1) that in the uncoupled model of Gao and colleagues similar changes in aperiodic/fractal components (H and 1/f slope) occur when the E:I ratio *g* is held constant, but the firing rate of E and I neurons is changed. In the uncoupled model, a decrease of H or the flattening of the 1/f slope could be due to an increase in the ratio of E:I conductance without changes in the firing rates, or to an increase in E firing rates over I firing rates without changes in ratio *g* of E:I conductance. Moreover, the uncoupled model does not capture the fact that E and I firing rates in cortex cannot be changed independently, as the coupling between E and I means that changing one of the two must change the other. Thus, these questions cannot be addressed with an uncoupled model.

### Supplementary Results: Using in-silico modeling to predict effects of chemogenetic perturbations of E:I balance on LFP or EEG biomarkers

To better understand the possible mechanisms behind the observed effects of the chemogenetic manipulations of E:I balance used in our three mouse experiments *in-vivo*, we simulated the *in-silico* applications of the 3 of chemogenetic manipulations. Results of our simulations are reported in Extended Data Fig. 3. We refer to the Methods section ‘*In-silico modeling of LFP and EEG data’*, for details of the simulation implementation.

The first *in-silico* manipulation mimicked the experimental effect of chemogenetic inhibition of both excitatory and inhibitory neurons via pan-neural hM4Di expression under the hSyn promoter. In this simulated manipulation, we decreased the excitability and firing of both E and I neurons (by decreasing the resting membrane potential, which makes it more difficult for neurons to fire given the level of input) and simultaneously weakened their synaptic strengths. This mimicks the effect of hM4Di activation, which primarily hyperpolarizes all neurons and secondarily suppresses synaptic release^26^. Our simulations predict that this manipulation would decrease excitability levels quantified by the average firing. The simulation also predicts that the decreased firing is accompanied by increased H, decreased 1/f slope, and decreased γ power measured from the LFP.

The second *in-silico* manipulation of the coupled E:I network mimicked the effect of experimentally-applied chemogenetic stimulation of pyramidal neurons via CamkIIα-promoter driven hM3Dq expression^26^. In this simulated manipulation, we increased the excitability and firing of the E neurons (by increasing the resting membrane potential) without altering their synaptic efficacies, to mimic the biological effect of hM3Dq receptor activation as reported in the experimental literature^26^. Our simulations predict that this manipulation would increase excitability levels quantified by the average firing, conformed by real data. The simulation also predicts that increased firing is accompanied by a decrease of H and an increase of 1/f slope, confirmed by real data. It also predicted a moderate (with respect to the next perturbation) increase in γ power. The latter could not be replicated in real data. One possibility is that CamkIIα exerts a secondary effect also on some inhibitory neurons, which has been reported by some studies^28^ but was not included in our simulations, and these secondary effects may affect gamma power. However, we verified that in this specific dataset the γ power had no additional predictive power on the firing rate with respect to H, in that a bilinear regression of firing rate against H + γ had the same performance of a regression based on H only). Thus, we did not explore in detail how to model the change of γ power in the CamkIIα dataset.

The third *in-silico* manipulation mimicked the effect of chemogenetic inhibition of PV neurons via cell-type specific expression of hM4Di receptors in this inhibitory population. In this simulation, we decreased the excitability and firing of I neurons (by decreasing the resting membrane potential) while simultaneously reducing their synapses conductances. This is done to reproduce the biological effect of hM4Di receptor activations, which is to hyperpolarize PV neurons and to decrease the release probability of the synapses they make. Our simulations predict that this manipulation would increase excitability levels quantified by the average firing. The simulations also predict that increased firing is accompanied by increased H and decreased 1/f slope and an increase of γ power observed in the LFP. The reason why γ power increases is that it arises from the engagement of the E-I loops. The perturbation potentiates this loop because E firing increases due to a decrease of I hyperpolarizing input from I to E neurons. This in turn produces a higher depolarizing input from E to I neurons that overcompensates for the higher threshold for firing of I neurons and leads I neurons to increase their activity.

As we do not know the exact changes of microscopic parameters induced in each experimental session by the chemogenetic perturbations, these simulations are not intended to reproduce the exact value of firing and LFP spectral changes found in the data. They also assume that activity at all frequencies are all recorded with the same SNR, which may not be true. However, these simulations provide a consistent direction of change (increase or decrease) of both firing rates and spectral features (H, 1/f slope, and γ power) across the entire explored range of variations in simulated microscopic parameters that we explored, a range that was sufficiently wide to contain the empirical effect sizes of changes in H and spectral features found in experimental values. These simulations provide a partial mechanistic explanation of why different ways to manipulate firing rate levels can lead to different changes in spectral features.

## Supplementary Figures

**Supplementary Fig. 1:**
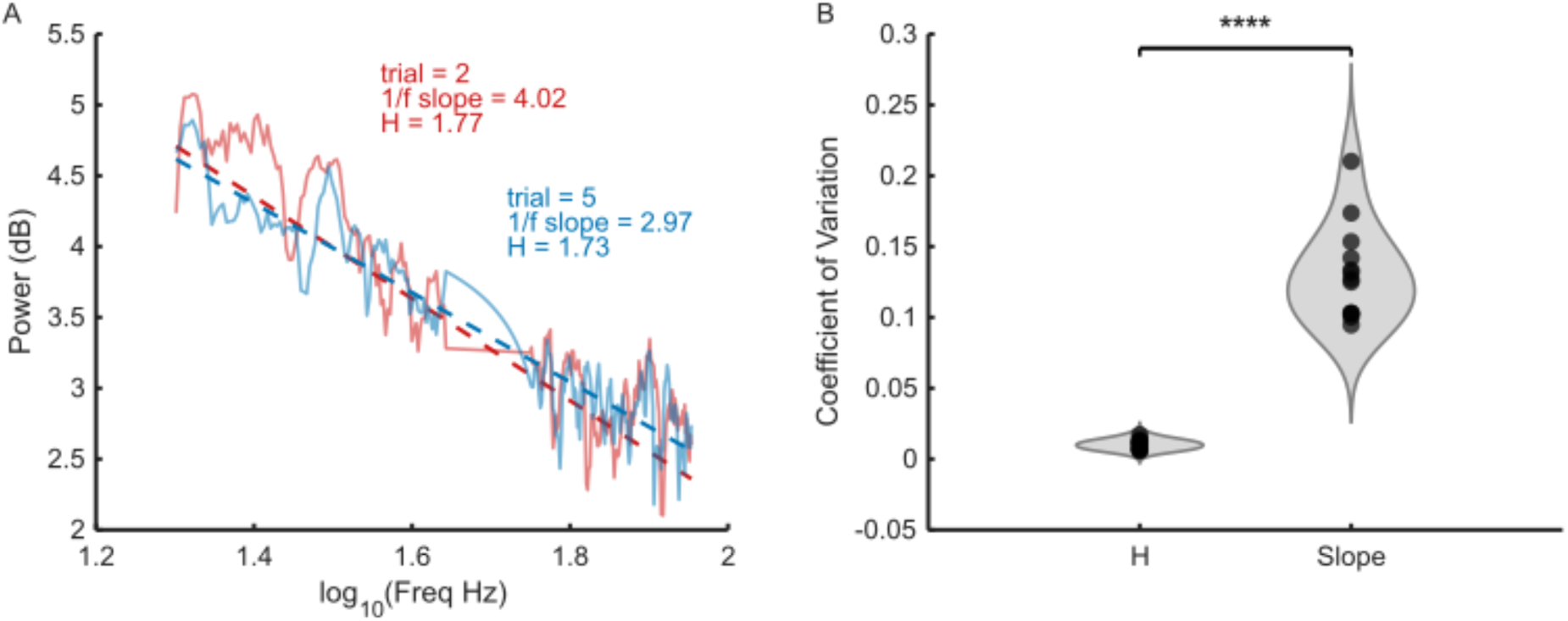
Variability of aperiodic spectral measures across trials. **a**, Example power spectral density plots (PSDs) from two non-consecutive trials within a 1-minute recording window of the CamkII-hM3D(Gq) experiment. This window was selected because the signal exhibits long autocorrelation (autocorrelation time ≈ 1 min), implying that consecutive PSDs, and their aperiodic components, should be similar (see methods). Solid curves show the raw PSDs, and dashed curves show the aperiodic fits obtained with the FOOOF algorithm. For each trial, both the 1/f slope and the Hurst exponent are displayed. Although the two PSDs are largely overlapping aside from stochastic fluctuations, the estimated 1/f slopes differ substantially, whereas H remains markedly more stable across trials. **b,** Plot showing coefficient of variation (CV) of the trial-wise 1/f slope and H across the same 1-minute window for CamkII-hM3D(Gq) experiment. Each point represents one subject. H shows significantly lower variability than the 1/f slope (paired t-test), indicating that H provides a more stable characterization of the fractal component of the time-series under these recording conditions.

**Supplementary Fig. 2:**
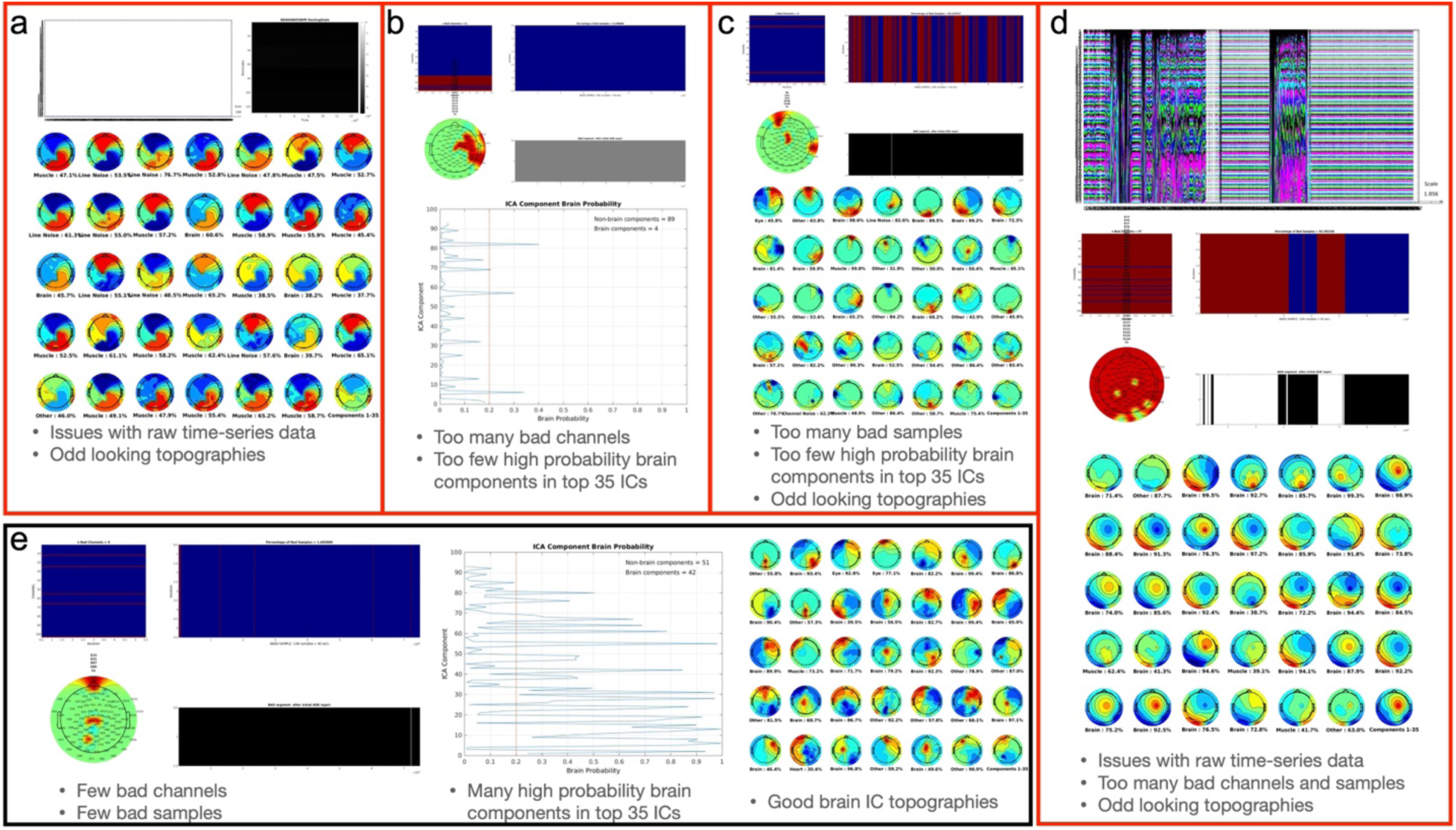
Examples of issues flagged up in data quality control analysis that warranted exclusion or inclusion of data into further downstream analysis. **a-d**, Plots that show different examples of issues identified in data quality inspection at different step of the preprocessing pipeline. **e,** Example participant with relatively clean data and which was included in further downstream analysis.

**Supplementary Fig. 3:**
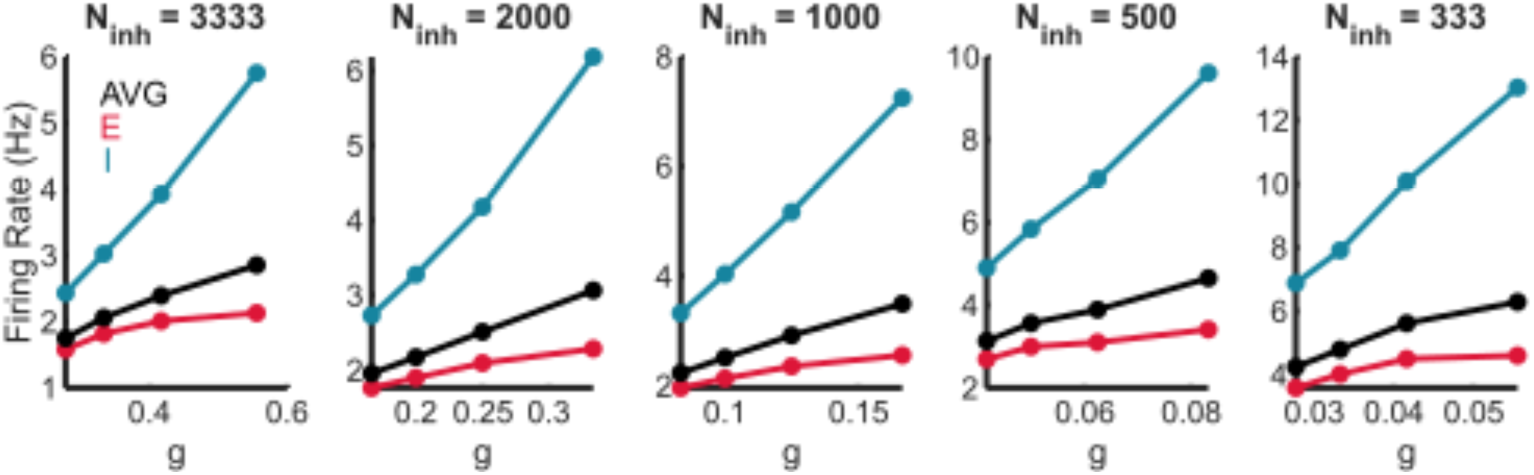
Simulations with different numerosity of inhibitory neurons. Firing rates of E and I neuronal populations, together with their weighted population-average, across simulations in which the number of inhibitory neurons was systematically varied (from highest to lowest, left to right). Synaptic weights were adjusted in each condition to maintain the same balance of excitatory and inhibitory input currents at the single neuron level as in the default network model used for the main figures

**Supplementary Fig. 4.**
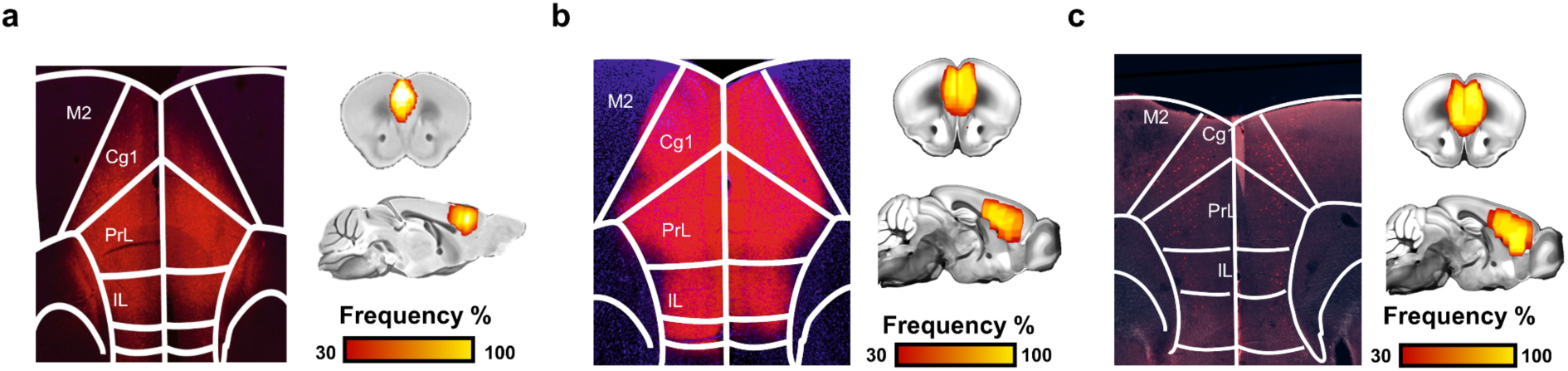
Viral expression localization. DREADD receptors were transduced bilaterally into the PFC of adult male mice. In each panel, the visualization on the left is a representative histology sample showing DREADD expression in red. The visualizations on the right of each panel are heatmaps that illustrate a qualitative regional assessment of viral expression across subjects. Panel **a** represents the hSYN-hM4Di experiment. Panel **b** represents the CamkII-hM3Dq experiment. Panel **c** represents the PV-hM4Di experiment.

## Supplementary Table Legends

**Supplementary Table 1:** This table reports the Pearson’s r correlations between network parameters (g, ν) and spectral features (H, 1/f slope, total power, γ) in the in-silico dataset, obtained from the first set of network simulations (i.e. Fig. 1). Top row shows correlations for LFP while bottom row shows correlations for EEG proxies (as shown in Fig. 1e-f).

**Supplementary Table 2:** This table provides the predictive performance of regression models used to estimate FR in the in-silico data (i.e. Fig. 1, Extended Data Fig. 2d). Model performance was assessed using 5-fold cross-validation, and the table reports Pearson correlation coefficients (mean ± 95% CI) between predicted and actual FR across folds. Analyses were performed on both LFP- and EEG-derived simulated signals, with each data point corresponding to a single simulation.

**Supplementary Table 3:** This table provides the results of linear mixed-effects models performed on in-vivo data for each chemogenetic experiment (hSyn-hM4Di, CaMKIIa-hM3D(Gq), PV-hM4Di). The statistical model is shown in the column labeled ‘model’. Each model had a different dependent variable. These models go along with the results shown in Fig. 2 and Extended Data Fig. 4. The table reports the F statistic, degrees of freedom, p-value, and partial η² for the group*condition interaction.

**Supplementary Table 4:** This table reports Pearson’s correlation between the actual firing rates and the model’s predicted firing rates (predictions over 10-fold cross-validation) for the 3 chemogenetic experiments. In the ‘Model’ column we specify the model used to predict firing rates (e.g., FR_pred_H+γ is the predicting FR from a model using H and γ as predictors). These statistics go along with the results shown in Fig. 2d, f, and h, and Extended Data Fig. 4. Statistics are reported for both SHAM and DREADD conditions.

**Supplementary Table 5. Predictive performance and statistical comparison of multi-feature models in vivo.** The table reports Pearson’s r correlations and 95% CI (first 2 rows) for the association between the actual firing rate and the predicted firing rates of different models. Each model predicting firing rates starts with H + γ (e.g., column 2), and subsequently adds another frequency band as a predictor (i.e. columns 3-6 add δ, θ, α, or β). Row 3 reports p-values from paired statistical tests performed across the 10 cross-validation folds, comparing each extended model against the H + γ baseline model. Non-significant p-values indicate that the model is not significantly different when compared to the model using H+γ as predictors.

**Supplementary Table 6:** This table provides the predictive performance of regression models used to estimate FR in the chemogenetic experiments (Fig. 2k). Model performance was assessed using 10-fold cross-validation, and the table reports Pearson correlation coefficients (mean ± SEM) between predicted and actual FR across folds. All models are trained and evaluated across pooled trials from the treatment phase of the experiments (hSyn-hM4Di, CaMKIIa-hM3D(Gq), and PV-hM4DiV), with each data point representing a single trial. The last column (i.e. FR ∼ H + γ (simulation weights)) uses beta weights for H and γ from the regression model on in-silico data rather than betas computed from real in-vivo experimental data.

**Supplementary Table 7:** This table shows model comparisons between the two models indicated in the first two columns. Each model tries to predict the FR using different sets of predictor variables (e.g., 1/f slope, total power, H, γ, H + γ, etc.). The z-statistic and p-values reported in columns 3 and 4 are derived from inferential statistical tests comparing model performance. Specifically, for each model, predictive performance was estimated using 1000 bootstrap resamples of the data, and the mean Pearson correlation coefficient between predicted and observed FR was computed across bootstrap iterations. Pairwise comparisons between models were then performed using two-sided z-tests on the difference between bootstrap mean correlations, with the pooled standard error estimated from the standard deviations of the corresponding bootstrap distributions. The null hypothesis is that both models are equally powerful at predicting variance in firing rates; p < 0.05 indicates that one model significantly outperforms the other. The sign of the z-statistic indicates which model performs better (a positive sign indicates the first model is superior, whereas a negative sign indicates the second model performs better). All models are trained and evaluated on pooled single-trial data from the treatment phase of the hSyn-hM4Di, CaMKIIα-hM3D(Gq), and PV-hM4Di experiments.

**Supplementary Table 8**: This table reports the results of all reval and SigClust analyses on the CMI-HBN dataset. In these analyses we ran autism-only or TD-only analyses on eyes open (EO) or eyes closed (EC) data. We re-ran the analyses 20 times with different random seed states for partitioning the data into training and validation sets. The analyses were also run with two different clustering algorithms (e.g., k-means or agglomerative hierarchical clustering) and with two different UMAP nearest neighbor values (e.g., n_neighbors = 30 or 5). Finally, SigClust p-values are reported on both training and validation sets for each analysis.

**Supplementary Table 9**: This table reports all statistics from EEG analyses of group differences in the CMI-HBN dataset. The columns are labeled according to the analysis that was implemented (e.g., CMI EO H, CMI EC H, CMI EO γ, CMI EC γ). The rows indicate each EEG electrode testing, while columns report F stats and p-values for subtype, age, and the subtype*age interaction. Follow-up t-statistics, p-values, FDR, and Cohen’s d are reported for TD vs A1 and TD vs A2 comparisons.

**Supplementary Table 10**: This table reports all behavioral variables analyzed from the CMI-HBN dataset. The first set of columns highlighted in yellow describe each of the variables and which domain they belong to. The variables are also labeled according to ‘result_type’ to indicate the type of difference was found, if any. The remaining columns report t-stat, p-value, Cohen’s d, and FDR values for each of the 3 pairwise group comparisons (e.g., TD vs A1; TD vs A2, A1 vs A2).

**Supplementary Table 11**: This table reports statistics for all PLS analyses. The first column labels each row according to the latent variable (LV) pair. Column 2 (eye_cond) labels the analysis by eyes open (EO) or eyes closed (EC). The 3rd column (DV) indicates where the brain data used was H or γ. The 4th column reports the PLS singular value (δ) for each LV pair. The 5th column reports the percentage of covariance explained for each LV pair. Columns 5 and 6 report the p-value and FDR value for each LV pair.

**Supplementary Table 12**: This table shows descriptive statistics for the CMI-HBN and IIT LAND datasets. The table breaks down sample sizes, age, and full-scale IQ for the EEG, behavioral, and PLS analyses.

**Supplementary Table 13**: This table provides a comprehensive description of the network model architecture and simulation settings, including population structure, connectivity, neuron and synapse models, and global simulation parameters used in the in vitro simulations.

**Supplementary Table 14:** This table provides a detailed summary of the neuronal and synaptic biophysical parameters, including membrane properties, synaptic kinetics, conductances, and external input statistics across all simulation sets.

